# Exome-wide rare variant genetic load is associated with cognition, intellectual disability and psychiatric disorders in population-based cohorts

**DOI:** 10.64898/2026.10.13.26365349

**Authors:** Mohammad Ahangari, Spencer Moore, Justin Schleede, Jonathan Anomaly, Alexander Strudwick Young, Tobias Wolfram

## Abstract

Rare protein-truncating variants (PTVs) and damaging missense variants in constrained genes are enriched among people with autism spectrum disorder (ASD), schizophrenia and related conditions, but most studies used clinical cohorts. In European-ancestry participants of the Adolescent Brain Cognitive Development Study (ABCD, 3,523 children) and the All of Us Research Program (AoU, 302,421 adults), we summed, separately for rare PTVs and damaging missense variants, the selection coefficients of affected genes into genetic loads (GLs). In ABCD, one additional typical constrained gene with a PTV was associated with 0.22 SD lower cognition and with ASD (odds ratio 1.80). In AoU, it was associated with ASD (1.34), schizophrenia (1.51) and intellectual disability (ID, 2.02). Missense GL showed smaller, concordant associations. Risk rose with gene constraint, to almost ninefold odds of ID in the most constrained genes, and carriers of a constrained PTV (12% of AoU participants) made up 31% of ID, 20% of schizophrenia and 17% of ASD cases. GL was essentially uncorrelated with polygenic scores. In African-ancestry participants, PTV GL retained 71–125% of its European-ancestry effect, versus 23–62% for matched polygenic scores. Constraint-weighted rare variant load contributes measurably to cognitive and neuropsychiatric outcomes in population cohorts.

## 1 Introduction

Cognitive ability, intellectual disability (ID) and neuropsychiatric conditions such as autism spectrum disorder (ASD) and schizophrenia have complex genetic architectures that are partially shared^1^. Large sequencing studies have shown that both rare protein-truncating variants (PTVs) and damaging missense variants are more common in people with ASD and schizophrenia, particularly when they affect genes that are evolutionarily constrained, i.e. depleted of functional variation in the general population^2–7^. Both *de novo* and rare inherited coding variants contribute to ASD and schizophrenia risk, with effects ranging from large to modest^4,5,7,8^. These findings suggest the impact of rare variants on ASD risk lies on a continuum between highly penetrant molecular diagnoses and modest, incompletely penetrant risk alleles^5,6^.

Evolutionary constraint provides a promising metric to estimate the extent to which damaging variants in a given gene are likely to increase risk of poor neurodevelopmental outcomes. Damaging variants arise continually by mutation and are removed by purifying selection, and the balance between the two maintains a mutational load of rare variants in every population^9^. The heterozygous selection coefficient (sHet), estimated from the depletion of PTVs in large sequencing datasets, quantifies the fitness cost of losing one gene copy, and the more constrained a gene, the more recent and rarer its damaging variants^2,10–13^. Aggregating rare damaging variants across constrained genes, or weighting them by sHet, has therefore been used to summarize their burden^14–16^. Because sHet approximates the fitness cost of heterozygous loss of function, we refer to the sHet-weighted sum as exome-wide rare variant genetic load (GL). The phenotypic consequences of GL calculated in this way appear to be wide-ranging. For example, it has been previously reported that damaging variants in evolutionarily constrained genes are associated with increased risk of ASD, attention-deficit/hyperactivity disorder (ADHD), schizophrenia, and other severe developmental disorders^4–7,14,17,18^. In adults, GL has been shown to be associated with lower cognitive performance and educational attainment, reduced height, and a wide range of fitness-related outcomes^14,15,19–21^. Most recently, scores summing gene-level selection coefficients over rare damaging variants have been related to cognitive performance across development, to educational outcomes, and to emotional and behavioral symptoms in British birth cohorts^22–24^.

Most rare variant studies of ASD and schizophrenia have been designed for gene discovery in clinically ascertained cases or families^3–5,7^, and population studies of recorded diagnoses, cognition and socioeconomic outcomes have mostly used binary sets of constrained or curated genes^14,19,25,26^. Less is known about whether a continuous, constraint-weighted GL is associated with ASD, schizophrenia and other recorded diagnoses in population cohorts of children and adults. If constraint indexes the expected severity of heterozygous loss of function, risk should increase with the constraint of the affected gene and with the number of constrained genes affected, rather than being confined to genes above a threshold. Established neurodevelopmental and schizophrenia genes, such as those curated in the Developmental Disorders Genotype-to-Phenotype database (DDG2P)^27^, the Simons Foundation Autism Research Initiative (SFARI) Gene database^28^ and the Schizophrenia Exome Sequencing Meta-analysis (SCHEMA) consortium^7^, account for only part of the rare variant contribution to these conditions^4,5,7^, and it is unclear how much constraint-weighted load lies outside them.

Rare damaging variants and common-variant polygenic scores (PGS) appear to contribute largely additively to neurodevelopmental conditions^6,16^, but whether this holds for a constraint-weighted load across cognitive and psychiatric outcomes in population cohorts is less clear. The two measures may also differ in how well they transfer across ancestries: PGS derived mainly from European-ancestry genome-wide association studies (GWAS) are markedly less accurate in other ancestries^29^, whereas the schizophrenia risk conferred by rare PTVs in constrained genes appears to be conserved across populations^30^. Rare variant associations are also not immune to confounding by population structure and other between-family effects^31^. Because siblings inherit different subsets of their parents’ rare variants, a comparison of population and within-sibship estimates can indicate whether associations reflect such confounding.

Here, we used two population cohorts, one of children and one of adults: the Adolescent Brain Cognitive Development (ABCD) Study^32^ and the All of Us Research Program (AoU)^33,34^. We tested whether GLs of PTVs and missense variants were associated with ASD, schizophrenia and cognition. We hypothesized that both variant classes would show concordant associations with cognition, ASD, and schizophrenia, with larger effects for PTVs. We then examined other psychiatric outcomes and height as exploratory analyses. In AoU, we additionally examined intellectual disability recorded in electronic health records (EHRs), as well as how associations varied with the constraint of the affected gene and with the number of constrained genes affected. We also examined the share of cases carrying a constrained PTV and the role of established disease genes. Finally, we examined the relationship of GL to PGS for the same outcomes, the consistency of associations across genetic ancestry groups, and associations within sibships. Primary analyses were restricted to European-ancestry participants for comparability between cohorts and with the available PGS. Other ancestry groups were analyzed separately.

## 2 Results

### 2.1 Constraint-weighted rare variant load in children and adults

We analyzed 3,523 genetically inferred European-ancestry participants in the ABCD cohort and 302,421 in AoU (Methods). In ABCD, NIH Toolbox total cognition was available for 3,498 participants^35,36^, and 3,506 participants had parent-reported ASD status, including 46 cases and 3,460 controls. In AoU, we defined ASD from EHR diagnoses or self-report, identifying 2,797 cases and 185,392 controls (*n* = 188,189). A GWAS of the AoU ASD phenotype gave a genetic correlation of 0.88 (SE 0.16) with the ASD GWAS of Grove et al.^37^ (Methods). We quantified the GL of PTV and damaging missense variants separately, summing for each individual the number of genes affected by a given variant type, weighted by the gene’s sHet^13^. The PTV and missense GLs were not significantly correlated (r = 0.016 in ABCD, 95% CI −0.017 to 0.049, *P* = 0.35). In ABCD, PTV GL had a mean of 0.047 and an SD of 0.090, so that one typical impacted constrained gene with a PTV (0.221 units of GL) corresponded to 2.46 SD of PTV GL. Because GL is strongly right-skewed, this increment lay at about the 96th percentile (Supplementary Figure S1). In AoU, PTV GL had a mean of 0.062 and an SD of 0.103, so that the same increment corresponded to 2.14 SD, or approximately the 94th percentile, and the mean sHet of constrained PTVs (0.217) was close to the ABCD increment. In ABCD, 76% of PTVs and 57% of damaging missense in constrained genes (sHet > 0.1) were seen in only one participant and absent from gnomAD. We refer to a qualifying PTV in a gene with sHet > 0.1 as a constrained PTV, and to participants carrying at least one as carriers. Carriers made up 8.9% of ABCD participants and 12.0% of AoU participants. In ABCD, carriers were about twice as common among children of caregivers with at most a high-school education as among children of caregivers with a graduate degree (16% versus 8%, trend *P* = 0.005, Supplementary Figure S2).

### 2.2 PTV and missense load are independently associated with lower cognition in children

Higher PTV GL was associated with lower NIH Toolbox total cognition (*β* = −0.088 SD per SD of GL, *P* = 4.5×10^−9^, Table S1). PTV GL accounted for 0.760% incremental R^2^. Missense GL showed a smaller association (*β* = −0.044, *P* = 0.011), corresponding to 0.193% incremental R^2^. We then regressed cognition on the PTV and missense GLs jointly. The incremental R^2^ of the joint model was 0.941%, approximately the sum of the two marginal values (0.953%), and both coefficients were nearly unchanged in the joint model (Table S1), indicating that the two GLs capture largely non-overlapping signal. To make the associations more biologically interpretable, we rescaled the fitted coefficients to quantify the effect of having one additional damaging PTV or missense variant in a typical constrained gene (Methods). Under the fitted linear model, one typical constrained PTV was associated with a 0.218 SD reduction in total cognition (95% CI 0.145 to 0.291), whereas one typical constrained damaging missense variant was associated with a 0.074 SD reduction (95% CI 0.017 to 0.130). On the standard-score scale (sample SD 16.3 points), these reductions corresponded to 3.5 points (95% CI 2.4 to 4.7) and 1.2 points (95% CI 0.3 to 2.1). A linear mixed model with a family random intercept, an alternative way of accounting for relatedness, gave a similar PTV estimate (0.194 SD reduction, 95% CI 0.113 to 0.275, Methods).

### 2.3 Associations with cognition and intellectual disability increase with gene constraint

To examine whether the association with cognition depended on how constrained the affected gene was, we grouped participants by the sHet of the most constrained PTV they carried. Mean NIH total cognition in ABCD was lower in participants whose most constrained PTV was in a more constrained gene, from 107.2 points in participants with no qualifying PTV to 96.4 points in those whose most constrained PTV was in a gene with sHet > 0.4 (Figure 1a). Mean scores were 106.4, 103.9 and 101.4 points in children carrying zero (*n* = 3,187), one (*n* = 294) and two or more (*n* = 17) constrained PTVs, respectively. Carriers of a constrained PTV had higher odds of scoring in the bottom 2.3% of the ABCD distribution (*≤* 76 points), the share of a normal distribution that lies more than 2 SD below the mean, the conventional threshold for intellectual disability (OR 2.14, 95% CI 1.13–4.08). Because ABCD excluded children with a diagnosis of ID or of moderate or severe ASD, this low-score group does not correspond to a clinical ID diagnosis.

**Figure 1:**
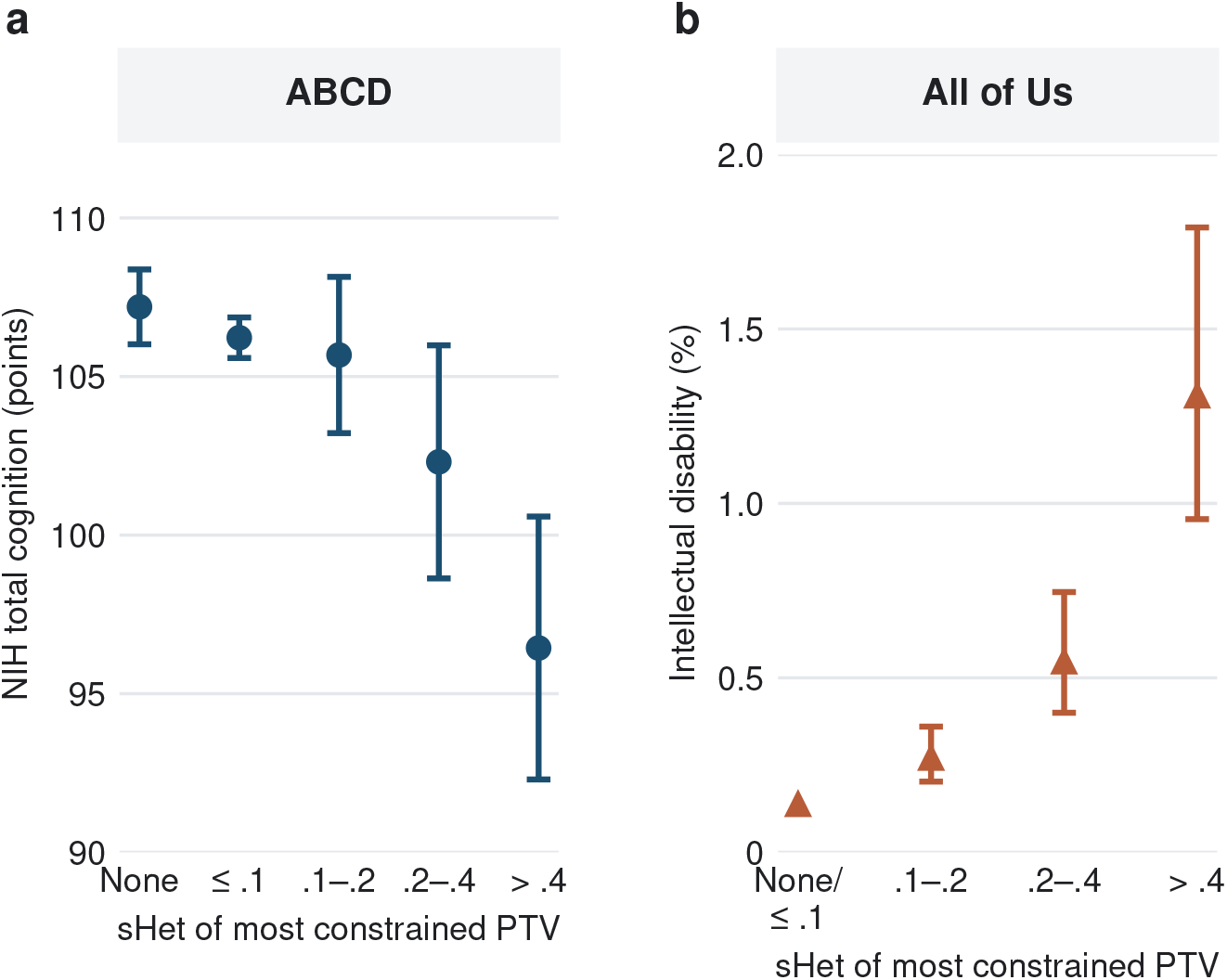
GL associations with cognition and intellectual disability by constraint group. (a) Unadjusted mean NIH Toolbox total cognition (age-corrected standard-score points) with 95% CIs in ABCD. Groups are participants with no qualifying PTV (*n* = 721), with PTVs only in genes with sHet *≤* 0.1 or without an sHet estimate (*n* = 2,466), or with the most constrained PTV in a gene with sHet 0.1–0.2 (*n* = 193), 0.2–0.4 (*n* = 84) or > 0.4 (*n* = 34). (b) Prevalence of EHR-recorded intellectual disability in AoU by the sHet of the most constrained PTV carried (0.1–0.2, 0.2–0.4 or > 0.4), with 95% Wilson CIs. The reference group combines participants with no qualifying PTV and those whose PTVs are only in genes with sHet *≤* 0.1.

We therefore examined EHR-recorded ID in AoU as an additional outcome (396 cases, *n* = 225,299). An additional typical impacted constrained gene was associated with ID for both PTV GL (OR 2.02, 95% CI 1.85–2.21, *P* = 4×10^−53^) and missense GL (OR 1.26, 95% CI 1.12–1.42, Table 1). ID prevalence increased with the sHet of the most constrained PTV carried, from 0.14% in participants without a constrained PTV to 1.31% in those whose most constrained PTV was in a gene with sHet > 0.4 (Figure 1b). Relative to non-carriers, the OR rose from 1.86 for sHet 0.1–0.2 to 8.85 (95% CI 6.29–12.44) for sHet > 0.4.

**Table 1:**
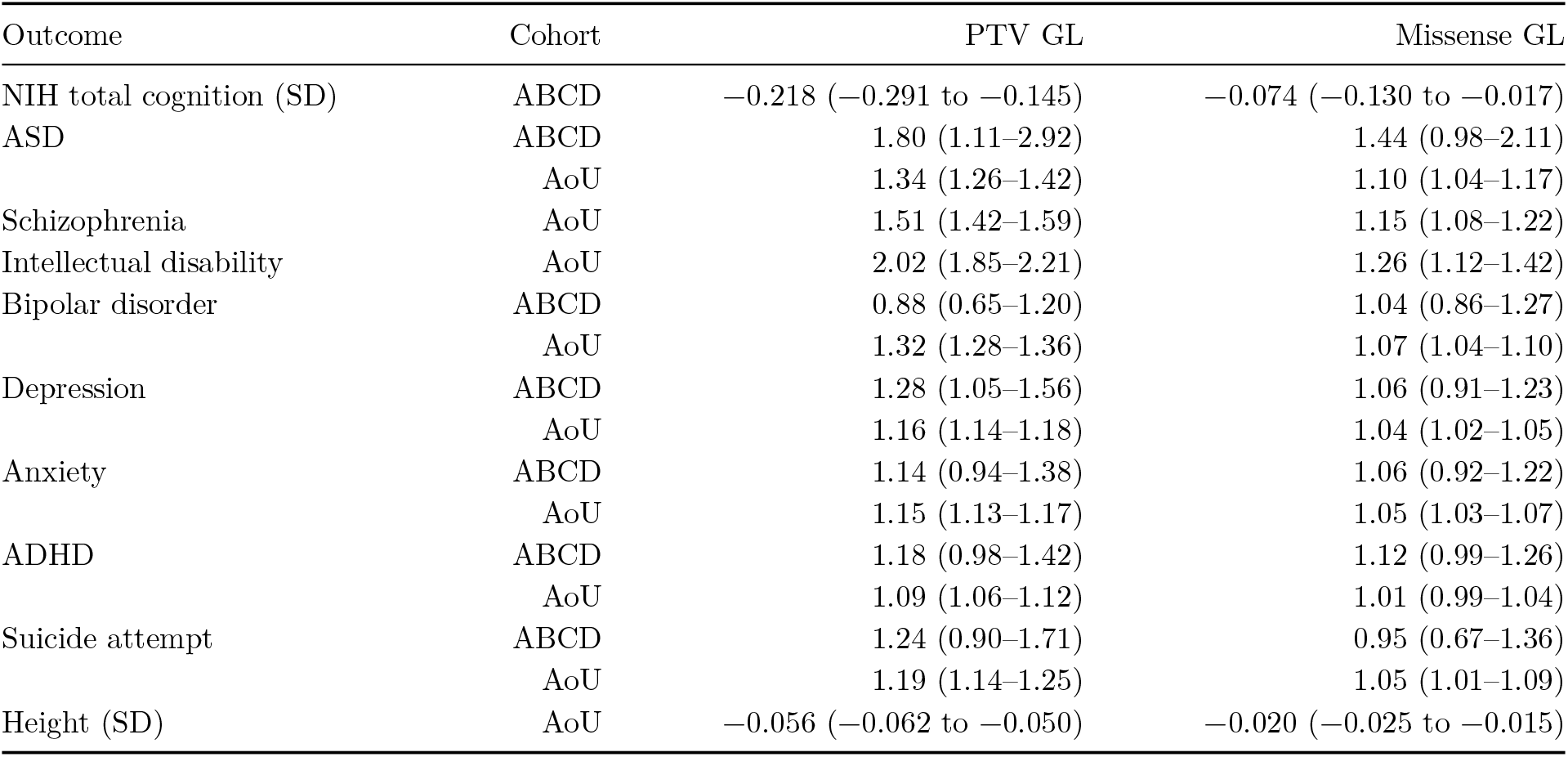
GL associations per typical impacted constrained gene. Values are odds ratios (ORs) for binary outcomes or SD differences for cognition and height, with 95% CIs, for PTV and damaging-missense GL in ABCD and AoU. Typical increments are 0.221 (PTV) and 0.259 (missense). Per-SD effects were converted to effects per typical impacted constrained gene with the GL SD of each analysis sample (Methods). These ORs are smaller than the ORs for carrying a constrained PTV in Table 2 (see Results).

### 2.4 PTV load is associated with ASD in both cohorts and with schizophrenia in adults

We then considered associations between the PTV and missense GLs and ASD in ABCD and AoU (Table 1, Figure 2a). ASD prevalence in the analyzed samples was 1.31% in ABCD and 1.49% in AoU. In ABCD, an increment corresponding to one typical impacted constrained gene was associated with an OR of 1.80 for PTV GL (95% CI 1.11–2.92, *P* = 0.017, Firth-penalized estimate 1.82) and 1.44 for missense GL (95% CI 0.98–2.11, *P* = 0.061). Marginal standardization^38^ of the ABCD models estimated that one constrained PTV increment above the sample-mean GL was associated with an increase in model-standardized ASD risk from 1.27% to 2.18%, corresponding to an adjusted risk ratio of 1.71 and an absolute increase of 0.91 percentage points. In AoU, the corresponding estimates, scaled using the ABCD-derived increment (Methods), were OR 1.34 for PTV load (95% CI 1.26–1.42, *P* = 7.8×10^−22^, EHR-only definition: OR 1.34) and OR 1.10 for missense load (95% CI 1.04–1.17, *P* = 0.0011), and the fixed-effect meta-analysis yielded ORs of 1.35 for PTV load (95% CI 1.27–1.43) and 1.11 for missense load (95% CI 1.05–1.18, Table S1). ABCD and AoU estimates were not significantly heterogeneous (*P* = 0.23 for PTV and *P* = 0.18 for missense). In AoU, an additional typical impacted constrained gene was associated with schizophrenia (2,306 cases and 186,703 controls, defined from EHR diagnoses or self-report) for both PTV GL (OR 1.51, 95% CI 1.42–1.59) and missense GL (OR 1.15, 95% CI 1.08–1.22). Schizophrenia was not analyzed in ABCD because of the limited case count.

**Figure 2:**
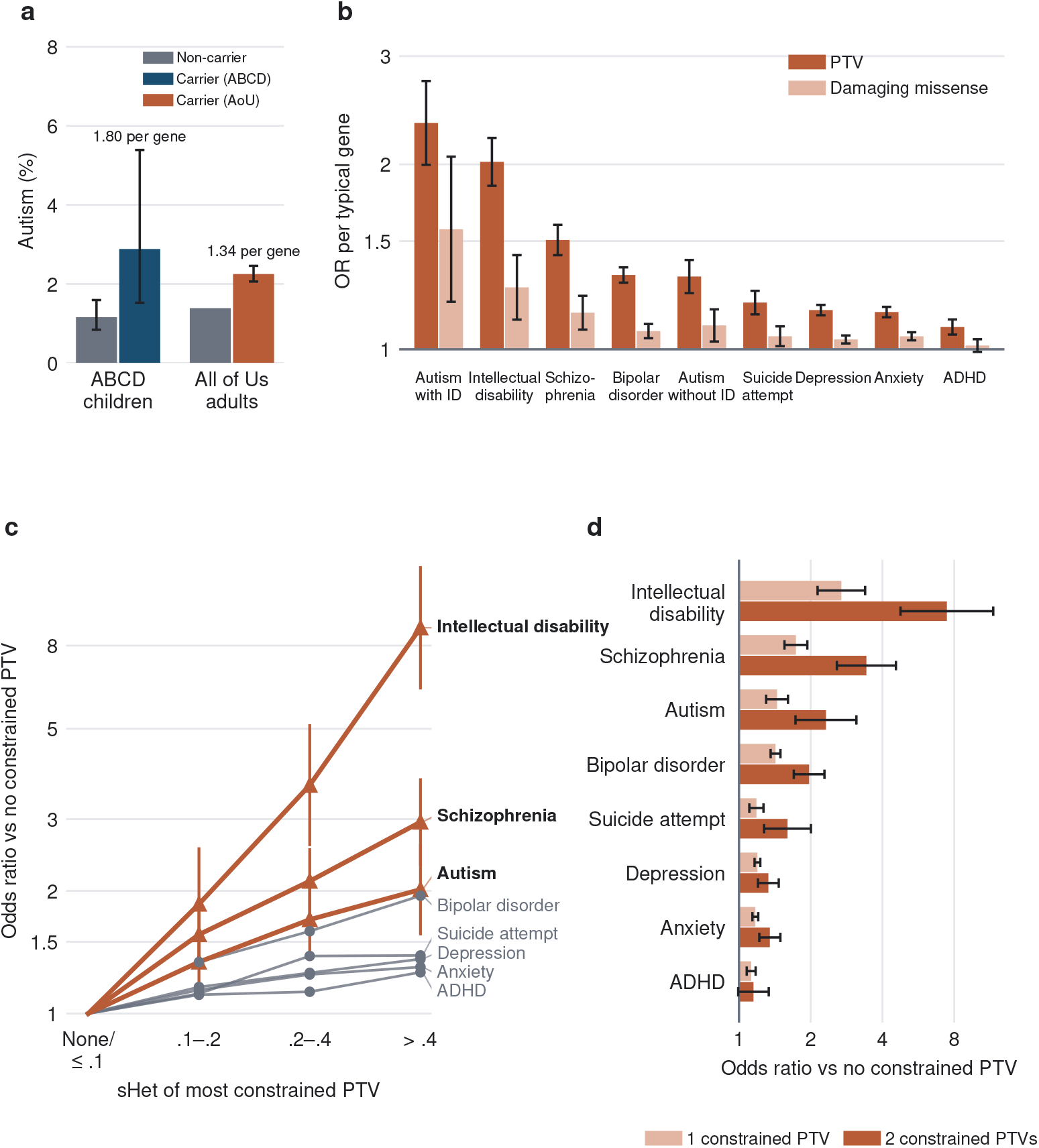
GL and neuropsychiatric traits. (a) ASD prevalence in non-carriers and carriers of a constrained PTV in ABCD (parent-reported ASD, 3,506 children, 46 cases) and AoU (EHR or self-reported ASD, 188,189 adults, 2,797 cases). Labels give the GL OR per typical impacted constrained gene (Table 1), not the carrier contrast shown by the bars. Error bars are 95% Wilson CIs. The AoU non-carrier prevalence is shown without an interval. (b) OR per typical impacted constrained gene in AoU for PTV and damaging-missense GL (scaled as in Table 1), ordered by PTV effect. ASD is split by co-occurring intellectual disability. (c) ORs relative to non-carriers by constraint group for eight outcomes. Intervals are shown for intellectual disability, schizophrenia and ASD. (d) ORs for carrying one or two constrained PTVs relative to none.

### 2.5 Associations extend to other psychiatric outcomes and rise with constraint and dose

We next examined exploratory associations with depression, broad anxiety disorder, bipolar disorder, ADHD, and suicide attempt. In AoU, higher PTV GL was associated with increased odds of all five outcomes, with estimated effects per typical impacted constrained gene ranging from OR 1.09 for ADHD to OR 1.32 for bipolar disorder (Table 1, Figure 2b). In ABCD, PTV GL was nominally associated with depression (OR 1.28, *P* = 0.012, *q* = 0.12), although this association did not survive correction for multiple testing, and the other estimates were imprecise, though directionally concordant with AoU for all traits except bipolar disorder (OR 0.88). Associations with missense GL were smaller, with the AoU associations with depression, broad anxiety, bipolar disorder, and suicide attempt reaching significance after false discovery rate (FDR) correction (*q* < 0.05), whereas the missense association with ADHD did not (*P* = 0.25). To assess whether GL associations extend beyond cognitive and psychiatric traits, we also considered height^14,19^. We found that one typical impacted constrained gene was associated in AoU with 0.056 SD lower height for PTV load and 0.020 SD lower height for damaging-missense load (both *P* < 10^−16^, Table 1).

In AoU, per-gene PTV GL associations were largest for the two outcomes involving intellectual impairment, ASD with co-occurring ID (OR 2.34, 95% CI 2.00–2.73) and ID, intermediate for ASD without ID (OR 1.31, 95% CI 1.23–1.40), bipolar disorder and schizophrenia, and smallest for suicide attempt, depression, anxiety and ADHD (Figure 2b). We then compared carriers with non-carriers, grouping carriers by the sHet of the most constrained PTV they carried (Figure 2c). For every outcome, the OR was largest in carriers whose most constrained PTV was in a gene with sHet > 0.4, but for the psychiatric outcomes the increase per unit of sHet was smaller in the most constrained genes than proportionality would predict (Supplementary Note 1). The gradient across the 0.1–0.2, 0.2–0.4 and > 0.4 groups was steepest for ID and schizophrenia, similar for ASD and bipolar disorder, and shallower for suicide attempt, depression, anxiety and ADHD. Carriers of two constrained PTVs had higher ORs than carriers of one for every outcome examined (Figure 2d), although the difference was small for ADHD.

### 2.6 Constrained PTVs account for up to a fifth of cases, about half outside known disease genes

Because per-gene ORs do not show how many affected individuals carry a constrained PTV, we next compared carrier frequencies in cases with those in the full samples. Compared with 12.0% of all AoU participants, the share of cases who carried a constrained PTV was higher for every outcome and was highest for the outcomes with the largest per-gene associations: 31.1% of participants with ID, 19.8% with schizophrenia and 17.3% with ASD (Figure 3a, Table 2). Assuming that these associations are causal, carrying a constrained PTV accounted for 21% of ID, 9% of schizophrenia and 6% of ASD cases in AoU, and for 1–5% of cases of the other outcomes. In ABCD, 19.6% of children with parent-reported ASD carried a constrained PTV, compared with 8.9% of all children, an attributable fraction of 13% (95% CI 5–23%), compared with 4% for total cognition in the bottom 10% (Figure 3a).

**Figure 3:**
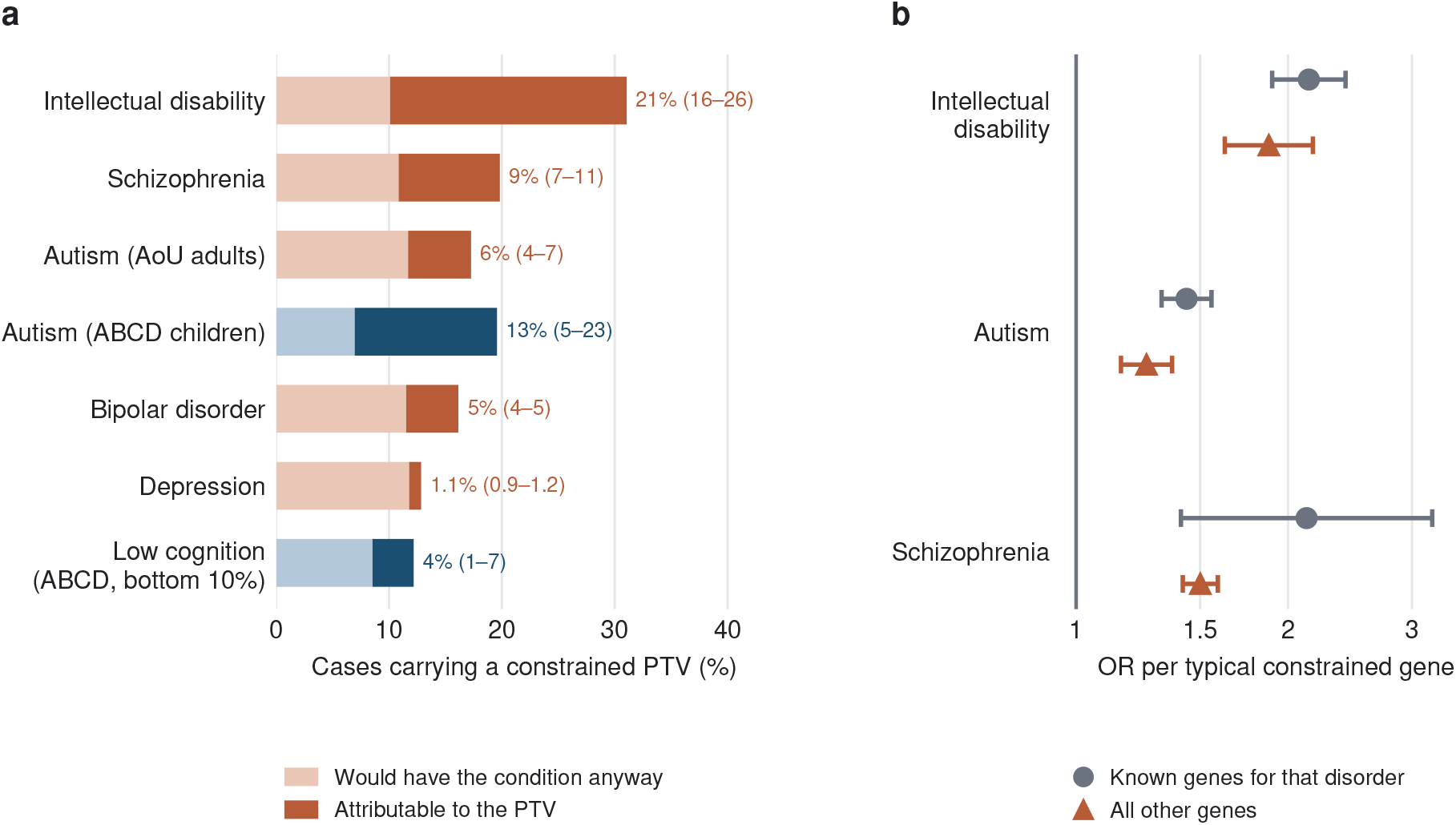
Constrained-PTV carriers among affected individuals, in and beyond known disease genes. (a) Percentage of cases who carry a constrained PTV in AoU (red) and ABCD (blue). The dark segment, labelled with its 95% CI, is the attributable fraction, the excess of carriers over those who would have had the condition anyway, assuming causal effects. (b) OR per typical impacted constrained gene in AoU, with PTV GL split into genes established for that disorder and all other genes. Established genes are DDG2P or SFARI genes for intellectual disability and ASD, and the nine autosomal SCHEMA exome-wide significant genes for schizophrenia. The wide interval for schizophrenia reflects the small number of genes.

**Table 2:** Constrained-PTV carrier status and neuropsychiatric outcomes in AoU. Carriers (10.9–12.0% of the analysis samples) carry at least one qualifying PTV in a gene with sHet > 0.1. Columns give the observed prevalence in non-carriers and carriers, the adjusted carrier OR, the percentage of cases who carry a constrained PTV (95% Wilson CI) and the attributable fraction (95% CI), the percentage of cases attributable to carrying a constrained PTV if the association is causal (Methods).

| Outcome | Prevalence,<br>non-carriers (%) | Prevalence,<br>carriers (%) | Carrier OR<br>(95% CI) | Cases who<br>carry (%) | Attributable<br>fraction (%) |
| --- | --- | --- | --- | --- | --- |
| Intellectual<br>disability | 0.14 | 0.45 | 3.09<br>(2.50–3.82) | 31.1<br>(26.7–35.8) | 21.0 (16.2–26.4) |
| Schizophrenia | 1.10 | 2.11 | 1.85<br>(1.67–2.06) | 19.8<br>(18.2–21.5) | 9.0 (7.4–10.9) |
| ASD | 1.39 | 2.25 | 1.51<br>(1.36–1.66) | 17.3<br>(15.9–18.7) | 5.6 (4.0–7.2) |
| Bipolar disorder | 7.83 | 11.53 | 1.47<br>(1.40–1.53) | 16.1<br>(15.6–16.7) | 4.6 (4.1–5.2) |
| Suicide attempt | 9.83 | 12.37 | 1.21<br>(1.13–1.29) | 13.4<br>(12.7–14.1) | 2.0 (1.4–2.7) |
| Depression | 49.54 | 54.80 | 1.21<br>(1.17–1.24) | 12.8<br>(12.6–13.0) | 1.1 (0.9–1.2) |
| Anxiety | 46.55 | 51.72 | 1.18<br>(1.15–1.22) | 12.9<br>(12.7–13.1) | 1.0 (0.8–1.1) |
| ADHD | 12.70 | 14.98 | 1.13<br>(1.09–1.18) | 13.2<br>(12.8–13.6) | 1.2 (0.8–1.7) |

Carrier ORs (Table 2) were larger than per-gene ORs (Table 1), for example 3.09 versus 2.02 for ID, although in ABCD carriers had on average only about one typical constrained gene more than non-carriers (Methods). Two features of the data explain this. First, risk did not rise in proportion to sHet (Supplementary Note 1). Per unit of sHet, effects were largest in moderately constrained genes, where most carriers have their PTVs, and smaller in the most constrained genes, whose large sHet values give them the most influence on the per-gene slope. Second, carriers are a mixture, and the minority with a PTV in a highly constrained gene had a much higher risk (for ID, OR 8.85), which raises the OR for carriers as a whole. The carrier OR therefore describes the risk of carrying a constrained PTV more directly than the per-gene OR, which summarizes the average gradient with GL.

Because the GL is based on evolutionary constraint rather than on established disease genes, we asked how much of the signal arises in genes already implicated in each disorder. In AoU, 79% of PTV GL lay outside genes listed in DDG2P^27^ or SFARI Gene^28^. When PTV GL was split into established genes and all other genes, the per-gene OR was larger in established genes for ID (2.14 vs 1.88, *P* = 0.18 for equal per-unit effects), for schizophrenia in the nine autosomal SCHEMA exome-wide significant genes^7^ (2.13 vs 1.50, *P* = 0.10) and for ASD (1.43 vs 1.26, *P* = 0.03), but the difference was nominally significant only for ASD. PTV GL outside established genes remained associated with each outcome (Figure 3b). GL excluding DDG2P and SFARI genes was associated with ASD (*P* = 5×10^−8^), and more strongly with schizophrenia and ID. Carriers whose constrained PTVs all lay outside established genes (DDG2P, SFARI or SCHEMA) accounted for 40% of the attributable fraction for ID, 50% for ASD and 61% for schizophrenia, and the SCHEMA genes for less than 1 of the 9 percentage points for schizophrenia. In ABCD, where established genes carried 18% of PTV GL, the association with cognition was similar for PTVs in established genes (−0.24 SD per typical gene, 95% CI −0.36 to −0.12) and in other genes (−0.21 SD, 95% CI −0.30 to −0.12, *P* = 0.73 for difference).

### 2.7 Rare variant load is essentially uncorrelated with and additive to polygenic scores

Because common variants also contribute to cognition and to each of these outcomes, we asked whether GL was correlated with trait-matched PGS and whether its associations were independent of them. In ABCD, PTV GL was weakly correlated with a cognitive PGS derived with SBayesRC^39^ (r = −0.035), and missense GL was not significantly correlated with it (r = −0.026). In the joint PTV and missense GL model, adjusting for the cognitive PGS did not significantly change the PTV GL association with cognition (from a 0.216 to a 0.195 SD reduction per typical impacted constrained gene, 95% CI 0.124 to 0.266) or the missense GL association (from a 0.072 to a 0.061 SD reduction). There was no evidence of a GL × PGS interaction (*P* = 0.23), and the ABCD ASD OR was essentially unchanged after adjustment for the ASD and cognitive PGS (1.82 before and 1.83 after adjustment in the joint model). Carriers of a constrained PTV scored lower than non-carriers within each cognitive-PGS tertile (by 1.6, 3.8 and 1.5 points in the low, middle and high tertiles).

In AoU, GL was essentially uncorrelated with each matched PGS (|r| *≤* 0.03), and adjusting for the matched PGS changed the PTV GL OR per typical impacted constrained gene by at most 2% (Figure 4a). We observed no GL × PGS interaction on the log-odds scale. When carrier status and PGS were considered jointly (Figure 4c), carriers in the lowest cognitive-PGS tertile had an OR for ID of 3.32 (95% CI 2.45–4.51) relative to non-carriers in the lowest tertile, whereas carriers in the highest cognitive-PGS tertile had an OR of 0.86 (95% CI 0.49–1.50) relative to the same reference group.

**Figure 4:**
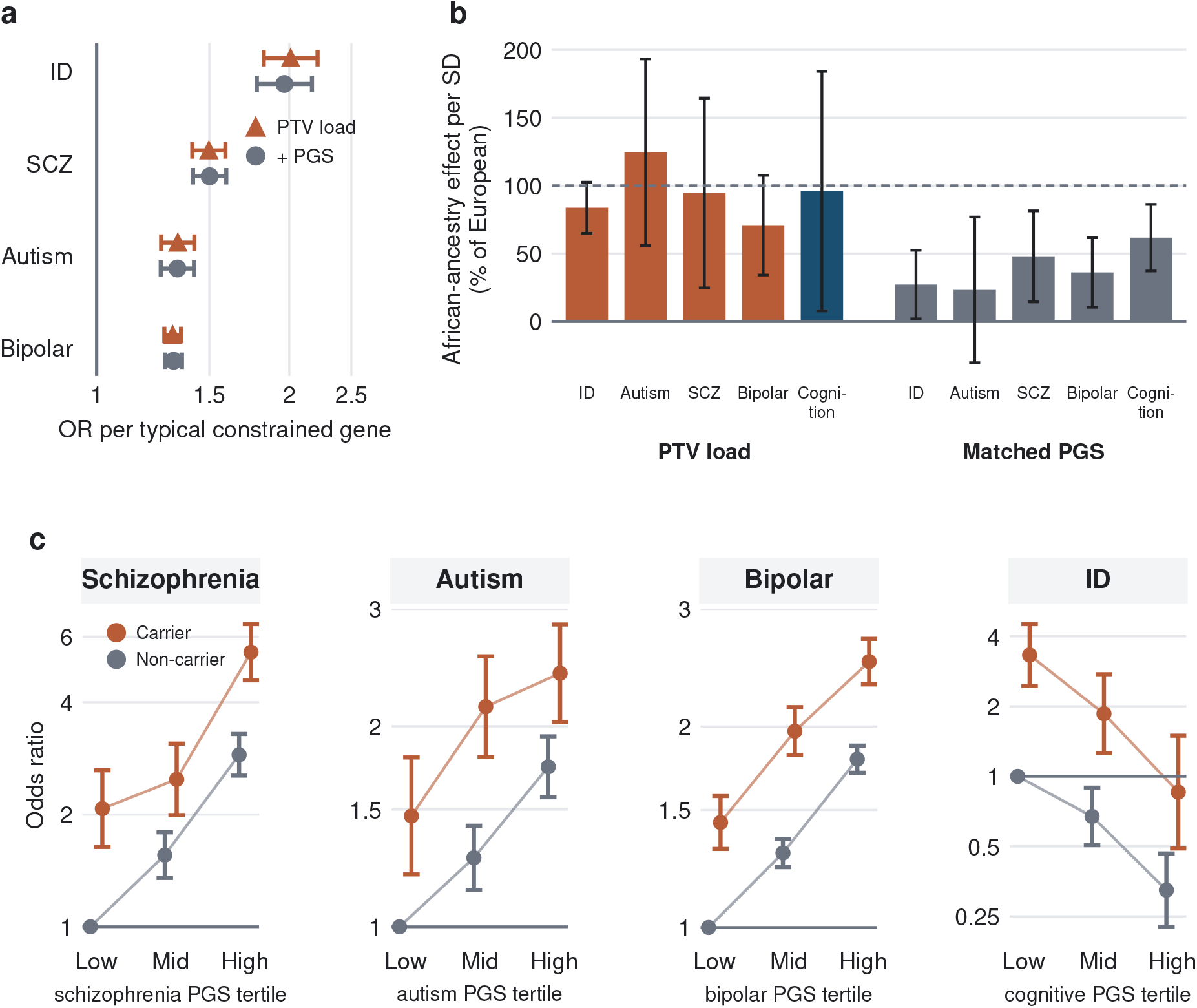
Rare-variant load and polygenic scores. (a) OR per typical impacted constrained gene for PTV GL in AoU, alone (triangles) and with the matched PGS (continuous) as a covariate (circles). The matched PGS is the cognitive PGS for intellectual disability and the disorder-specific PGS otherwise. (b) Effect in African-ancestry participants as a percentage of the effect in European-ancestry participants, for PTV GL and the matched PGS, both per SD within each ancestry group. Outcomes are intellectual disability, ASD, schizophrenia and bipolar disorder in AoU, and cognition in ABCD. The ABCD African-ancestry sample for the cognition comparison comprised 850 children. The dashed line marks equal effects. Schizophrenia and bipolar disorder use specific definitions for both PTV GL and the PGS: EHR diagnosis and self-report for schizophrenia, and self-report for bipolar disorder. Error bars are delta-method 95% CIs. (c) Carrier × PGS tertile in AoU for schizophrenia, ASD, bipolar disorder and intellectual disability, with carriers and non-carriers distinguished by color. ORs are relative to non-carriers in the lowest PGS tertile.

### 2.8 Rare variant load transfers across ancestries better than polygenic scores

To assess whether GL associations were consistent across ancestries and how their transferability compared with that of PGS, we analyzed all AoU genetic ancestry groups with a single reduced covariate set (Methods). The per-gene ASD association was consistent across ancestry groups (OR 1.34 in European-ancestry, 1.40 in African-ancestry, 1.36 in admixed American-ancestry, 1.33 in East Asian-ancestry and 1.59 in South Asian-ancestry participants, heterogeneity *P* = 0.97). Middle Eastern-ancestry participants had too few ASD cases to be analyzed. Expressed per SD and as a percentage of the effect in European-ancestry participants, the PTV GL effect in African-ancestry participants was 84% for ID (95% CI 65–103%) and 125% for ASD (95% CI 56–193%), compared with 27% (95% CI 2–52%) and 23% (95% CI −30 to 77%) for the matched PGS (Figure 4b). For schizophrenia and bipolar disorder, the comparison depended on the phenotype definition. PTV GL associations were attenuated in African-ancestry participants under EHR-based definitions but consistent across ancestries under more specific definitions, whereas the matched PGS remained attenuated (Figure 4b, Methods, Supplementary Note 2). In ABCD, the PTV GL association with cognition in African-ancestry children was 96% of that in European-ancestry children, compared with 62% for the cognitive PGS, close to the 66% reported for this PGS in African-American participants of ABCD^40^.

### 2.9 Within-family estimates are imprecise but do not differ significantly from population estimates

Because rare-variant associations can be affected by population stratification and other confounding^31^, we also estimated associations within sibships (Table S2). In AoU, within-sibship estimates from conditional logistic regression in 4,229 full-sibling pairs from 3,883 families (Supplementary Note 3) were imprecise but did not differ significantly from the population estimates for any outcome (all *P* > 0.15). For a combined ASD, schizophrenia or bipolar disorder outcome, the within-sibship OR was 1.83 (95% CI 1.11–3.01), compared with 1.27 in the population (*P* = 0.16 for the difference). ASD, schizophrenia and ID alone could not be estimated. In ABCD, the within-family estimate for cognition in 375 dizygotic-twin and sibling families was −0.085 SD per typical impacted constrained gene and not significantly smaller than the population estimate of −0.218 SD (family-cluster bootstrap *P* = 0.16), but imprecise (95% CI −0.255 to 0.085).

## 3 Discussion

We found that exome-wide GLs of rare protein-altering variants were associated with both lower cognition in ABCD and elevated risk of ASD, schizophrenia, EHR-recorded ID and other psychiatric conditions. PTV GL in general displayed stronger associations than missense GL, consistent with prior studies^3,4,7^, and joint modeling of cognition indicated that the two GLs were associated with it largely independently. The association with lower cognition confirms reports from British birth cohorts that used similar constraint-weighted scores^22^ and from studies of educational attainment^21^ and adult cognitive function^20^. Consistent with these reports, associations were graded by the constraint and number of affected genes, persisted in genes outside established disease gene lists and were additive to common-variant PGS, and we extend these findings to recorded psychiatric diagnoses in children and adults.

The constraint gradient was steepest for ID, for which carriers of a PTV in the most constrained genes had almost nine times the odds of non-carriers. It was also clear for schizophrenia, ASD and bipolar disorder but shallow for the other psychiatric outcomes, and the largest per-gene associations were for outcomes involving intellectual impairment, as in previous studies of rare damaging variants^14,17^. These patterns extend findings for multiple rare variants in dominant developmental disorder genes^25^ and for loss-of-function variants in curated neurodevelopmental genes^26^ to an exome-wide, constraint-weighted score. In AoU, however, the increase was less than proportional to sHet in the most constrained genes, significantly for most psychiatric outcomes and non-significantly for ID (Supplementary Note 1). Proportionality is not necessarily expected for any single outcome, because sHet indexes the total fitness cost of heterozygous loss of function rather than its effect on a specific trait.

GLs were weighted by an evolutionary constraint metric and therefore do not depend on whether a variant occurs in genes currently established as associated with cognition or ASD. Although constraint is estimated from population variation without reference to phenotype, the selection it reflects acts on the consequences of loss of function, among which impaired cognition and neurodevelopmental conditions are prominent^14,15^, so genes under strong selection are expected to be enriched for effects on these outcomes. This approach is thus able to capture GL in genes not yet well characterized in the clinical genetic setting as disease causing, potentially due to incomplete penetrance^19^. Most PTV GL lay outside established genes, and GL outside DDG2P and SFARI genes remained associated with ASD, schizophrenia and ID. Both observations agree with reports that much of the inherited rare variant contribution to ASD lies outside currently known genes^5^. Per unit of constraint, associations tended to be slightly larger in established genes, although the difference was nominally significant only for ASD. About half of the attributable fraction came from genes outside established lists. The attributable fraction for ASD in AoU (6%) was close to the 6% reported for rare inherited loss-of-function variants in a large family-based study^5^, although our estimate also counts *de novo* PTVs. It was about twice as high, though imprecise, in ABCD children. For schizophrenia, which could be examined only in AoU, the association was consistent with the excess of rare PTVs in constrained genes reported in case-control exome studies^7^ and was not confined to the SCHEMA genes.

Rare and common variation appear to capture largely distinct components of liability. GL was essentially uncorrelated with the PGS for the same outcomes, and the associations were approximately additive on the log-odds scale, as reported for ASD^6^, developmental disorders^16^ and educational attainment^25^. Schizophrenia risk was highest in carriers with a high schizophrenia PGS, and ID risk in carriers with a low cognitive PGS (Figure 4c).

The two measures also differed across ancestries. The GL association with ASD was consistent across five ancestry groups. In African-ancestry participants, the GL associations with ASD and cognition did not differ significantly from those in European-ancestry participants and the association with ID was only modestly smaller (84% of the European-ancestry effect per SD), whereas the matched PGS associations were significantly attenuated (27% for ID). For outcomes other than ID, the rare variant intervals were wide. The weaker schizophrenia and bipolar disorder associations in African-ancestry participants under EHR-based definitions were no longer significantly heterogeneous under more specific definitions, whereas the PGS remained attenuated, suggesting that this attenuation largely reflected phenotyping rather than differences in rare variant effects. Because selection removes variants in constrained genes within a few generations, most arose after the divergence of continental populations, and the deleterious load per genome differs little between populations^41,42^. Such variants are also assayed directly by sequencing rather than tagged through linkage disequilibrium. Together with the conservation of rare PTV effects on schizophrenia across populations^30^ and the known loss of PGS accuracy with genetic distance from GWAS discovery samples^29,40^, these results suggest that rare variant effects on a constraint scale transfer across ancestries more readily than PGS, although this needs confirmation in larger non-European samples.

The one primary outcome that could be tested within families was cognition in ABCD, and its within-family estimate was not significantly smaller than the population estimate but imprecise and compatible with anything from no to substantial confounding. Stronger evidence on direct effects comes from trio analyses in British birth cohorts, which found direct effects of rare variant burden on cognition similar to population effects^22^. Our within-family analyses in ABCD and AoU are too imprecise to exclude confounding for specific outcomes.

Constrained PTVs were also concentrated at the lower end of the socioeconomic distribution, as expected if GL lowers educational attainment in the parental generation^21^, so the yield of genetic testing for these variants is highest in families with the fewest resources.

We observed a larger point estimate for ASD in ABCD than in AoU, but the difference was not statistically significant. Less specific ASD diagnoses recorded in adulthood, in participants mostly born before the broadening of ASD criteria, could bias the AoU estimate toward the null. The AoU estimates for ID are also likely to be conservative. AoU enrolls only adults who can consent for themselves, and ID was recorded in 0.18% of participants with EHR data, compared with a population prevalence of about 1%^43^. Recorded ID in AoU is therefore mostly mild, and severe ID, which more often has rare causes of large effect^44^, is largely missing. The association with ID and its attributable fraction of 21% probably underestimate those in the general population. Similarly, brief assessments such as the NIH Toolbox measure general cognitive ability with considerable error^40^, so the association of GL with the underlying ability, expressed in SDs, is probably larger than estimated here.

This study has several limitations. First, the analysis depended on parental reports, self-reports and EHRs rather than standardized diagnostic assessments, so diagnoses may be missed or mis-classified, and participation in AoU may be related to health and health-care use. The exploratory psychiatric phenotypes were also broadly defined. In ABCD, they were derived from the parent-reported computerized Kiddie Schedule for Affective Disorders and Schizophrenia (KSADS-COMP) rather than from clinical diagnoses and pooled present, past, remission, other specified and unspecified diagnoses across up to eight visits. In ABCD, 29% of children met this definition for ADHD, although only 6.3% had a present ADHD diagnosis at baseline, and 7.5% for bipolar disorder, half of them with only an unspecified or other specified diagnosis. Case fractions also exceeded population prevalence in AoU (for example, 50% for depression, 47% for anxiety, 13% for ADHD and 8.3% for bipolar disorder). This may explain why ADHD had the weakest association here, whereas clinically diagnosed ADHD carries a burden of rare PTVs similar to that of ASD^17^. Second, only cognition, ASD and schizophrenia were primary outcomes. FDR correction was applied within the exploratory outcomes, but the analyses of ID, constraint groups, the number of constrained PTVs, established genes, PGS, ancestry and sibships were not corrected for multiple testing. Third, PTV calls were imperfectly reproducible, particularly for frameshift indels in ABCD. Missed calls reduce power and per-SD effects but not per-unit effects, whereas false-positive calls attenuate associations, and the measured GL reliability implies that this attenuation is modest (Supplementary Note 3). Fourth, aggregate GL cannot identify whether an association is driven by many genes or a small number of influential variants, and therefore provides only an average effect across the genes and variants that contribute to GL. Fifth, GL included only autosomal single-nucleotide variants and short indels. Copy-number, structural and X-linked variants, which also contribute to these conditions, were not captured, so the attributable fractions understate the total contribution of rare variants. Because a deletion removes one copy of every gene it spans, deletions could be added to GL by summing sHet over the deleted genes, as gene constraint has been summed over copy-number variants to estimate their effects on cognition and ASD^45,46^. Duplications would require separate measures of sensitivity to increased gene dosage^47^.

Taken together, the results support a broad set of associations between exome-wide rare variant GL and cognitive performance, ASD, ID, and other psychiatric outcomes in childhood and adulthood. Constraint-weighted GL provides a gene-list-free summary of rare coding variation that complements PGS in population studies of cognitive and neuropsychiatric outcomes, including studies in ancestry groups for which PGS remain less accurate.

## 4 Methods

### 4.1 Study design and analysis cohort

We performed GL association analyses using whole-genome sequencing (WGS) and phenotype data from the Adolescent Brain Cognitive Development Study (ABCD)^32^ and the All of Us Research Program (AoU)^33,34^. In ABCD (release 7.0), we restricted analysis to genetically inferred European-ancestry participants with WGS of whole-blood samples, identified by projection onto the 1000 Genomes Phase 3 reference^48^ and a European-assignment probability cutoff of 0.90. The analytic sample consisted of 3,523 genetically inferred European ancestry participants. In AoU, GL association analyses were restricted to 302,714 participants assigned European ancestry by the AoU genetic ancestry classifier^34^. After excluding 293 participants with predefined sample quality issues, the eligible cohort comprised a maximum of 302,421 participants for the final analysis. The primary cohort in AoU included whole-blood and saliva samples, and related participants were retained, with relatedness accounted for using family-clustered standard errors. Sample sizes for individuals with nonmissing data per regression analysis are available in Table S1.

Primary analyses were restricted to European-ancestry participants because ABCD provides an adequately sized sample only in this group and because the GWAS underlying the PGS and the population-genetic resources used for variant filtering are predominantly of European ancestry. Additional analyses included all AoU genetic ancestry groups. In accordance with the AoU Data and Statistics Dissemination Policy, AoU counts of 1–19 participants, and counts from which such values could be derived, are not reported.

### 4.2 Primary and exploratory phenotypes

The two primary outcomes in ABCD were NIH Toolbox total cognition^35,36,49,50^ (the age-corrected Total Cognition Composite score, from the baseline visit where available), rank-based inverse-normal transformed among all sequenced participants and analyzed quantitatively, and parent-reported autism diagnosis (whether the child had been diagnosed with ASD, from the parent screening interview at study entry), analyzed as a binary outcome. Higher GL was hypothesized to predict lower cognition and higher autism risk. For each AoU psychiatric outcome, cases had a qualifying EHR code or reported the diagnosis for themselves in the AoU surveys, and controls had answered the surveys’ mental health condition question without reporting the diagnosis and had no qualifying code, with no requirement for EHR data. Participants without survey data could therefore be cases but not controls, so sample sizes differ by outcome (Table S1, Supplementary Note 2). In AoU, ASD cases had an ICD-10-CM code F84.0, F84.5 or F84.9 or an ICD-9-CM code 299.0, 299.8 or 299.9, or reported an ASD diagnosis in the AoU survey. For the EHR-only ASD definition used in the sensitivity analysis, controls were participants with EHR data and no qualifying code. Schizophrenia was defined from EHR diagnoses (ICD-10-CM F20 excluding F20.81, or ICD-9-CM 295 excluding 295.4 and 295.7) or from a “Self” answer to the Personal and Family Health History or Personal Medical History survey item “Including yourself, who in your family has had schizophrenia?”. Codes for schizophreniform, schizoaffective and other psychotic disorders neither defined cases nor excluded controls. The resulting prevalence of schizophrenia in AoU (1.22%) was at the upper end of population-based epidemiological estimates of lifetime prevalence^51,52^. Height was taken from all recorded AoU height measurements, including program physical measurements and EHR records. We retained adult measurements (age *≥* 18 years, valid measurement date, 130–215 cm inclusive), converted measurements recorded in inches to centimeters, and defined each participant’s height as the median of the retained measurements, with age at the median-dated measurement used as the adjustment age. Height was rank-based inverse-normal transformed before standardization.

For the genetic correlation with the ASD GWAS of Grove et al.^37^, we used AoU v9 WGS data from unrelated European-ancestry participants and two-step REGENIE^53^ with approximate Firth correction, adjusting for age, sex and ten genetic principal components. The genetic correlation was estimated with cross-trait LD score regression^54,55^ in GenomicSEM^56^, using HapMap3^57^ SNPs, European 1000 Genomes^48^ LD scores and freely estimated intercepts.

Exploratory psychiatric phenotypes were harmonized approximately across cohorts. In AoU, EHR diagnoses were defined by the following codes, and self-report contributed additional cases for depression, broad anxiety, bipolar disorder and ADHD, defined from “Self” answers to the corresponding items of the same survey question (answer concept IDs 1384656, 1384475, 1384669 and 1384510). Depression was defined by ICD-10-CM F32–F33 or ICD-9-CM 296.2, 296.3, 300.4, and 311. Broad anxiety was defined by ICD-10-CM F41.1, F41.8, and F41.9 or ICD-9-CM 300.00, 300.02, and 300.09. Bipolar disorder was defined by ICD-10-CM F30–F31 or ICD-9-CM 296.0–296.1 and 296.4–296.8. ADHD was defined by ICD-10-CM F90.x or ICD-9-CM 314.x. Suicide attempt was defined by ICD-10-CM X71–X83, T14.91, and specified intentional-self-harm codes within T36–T65 and T71, or ICD-9-CM E950–E959 (these codes capture intentional self-harm irrespective of suicidal intent), and self-reported suicide attempt came from the life-time suicide attempt item of the AoU Emotional Health History and Well-Being survey, which fewer participants completed. In ABCD, approximately corresponding phenotypes were derived from parent-reported KSADS-COMP^58,59^ modules (for anxiety, the generalized anxiety disorder module) across all available visits and included present, past, remission, other, and unspecified diagnostic fields where applicable. Participants were classified as cases if any qualifying field was positive and as controls if they had an observed negative response with no positive response. Skip and missing codes were treated as missing. The ABCD suicide attempt phenotype included actual, interrupted, and aborted attempts.

### 4.3 Intellectual disability and low cognition

In AoU, ID cases had an EHR record of ICD-10-CM F70–F79 or ICD-9-CM 317–319, and controls were participants with EHR data (any condition record) and no ID code, without a survey requirement. ASD cases were further split by co-occurring ID: ASD with ID comprised ASD cases with any ID code, ASD without ID comprised ASD cases with no ID code, and both were compared with the ASD controls who had no ID code. Because children with a diagnosis of ID were not eligible for ABCD, an ID diagnosis could not be analyzed in ABCD. We instead analyzed low NIH Toolbox total cognition, defined as scoring in the bottom 10% of the analysis sample (*≤* 86 age-corrected standard-score points) or in the bottom 2.3% (*≤* 76 points). The bottom 2.3% is the share scoring below 70 in a population with mean 100 and SD 15, and the cutoff is higher here because the sample scored above the test norms.

### 4.4 Sequence data preparation

Sequence data processing was performed on the GRCh38 reference assembly and restricted to autosomes in both cohorts. ABCD analyses used release 7.0 short-read whole-genome VCFs aligned to the GRCh38 no-alt reference genome that were normalized and decomposed into biallelic records. AoU analyses used version 9 short-read whole-genome sequence data from the exome-region Hail^60^ MatrixTable derived from the WGS callset, and multiallelic loci were split into biallelic representation within Hail while retaining the original alleles and alternate-allele indices.

Genotype-level filtering differed between cohorts. In ABCD, PTV calls required genotype quality (GQ) >20, read depth (DP) >20 for SNVs or >30 for non-SNVs, and alternate-allele balance of 0.30–0.70, and missense calls required GQ *≥*20, DP *≥*10 and alternate-allele balance of 0.20– 0.80. In AoU, PTV and missense calls required GQ >25, DP >10, and alternate-allele balance of 0.30–0.70.

### 4.5 Exome-wide rare variant genetic load construction

Candidate variants in both cohorts were annotated with VEP v114.2^61^, including LOFTEE^2^, CADD v1.7^62^, REVEL v1.3^63^ and SpliceAI^64^ annotations, and the final masks retained consequences on MANE Select transcripts^65^, with reference and alternative alleles of at most 5 bp. PTVs were stop-gained, frameshift and canonical splice-site variants with a high-confidence LOFTEE prediction, together with start-lost and stop-lost variants with CADD PHRED greater than 25 (stop-lost variants were included in the PTV class although they do not truncate the protein). Splice-site PTVs also required a SpliceAI maximum delta score greater than 0.8 for the same gene, and PTVs in the terminal exon or terminal intron were excluded. Single-nucleotide PTVs with a CADD score were required to have CADD PHRED greater than 25, whereas indels were not filtered on CADD. Missense variants were required to have a REVEL score greater than 0.70 and CADD PHRED greater than 25. We required all genotypes to be heterozygous to reduce false positive calls. We required variants to have gnomAD v4.1.1^2^ global exome and global genome allele frequencies below 1×10^−5^, retaining variants absent from gnomAD, and in ABCD, missense variants also had to have non-Finnish European exome and genome allele frequencies below 1×10^−5^. We further required that variants be carried by at most eight individuals in each cohort (in ABCD, counted among the 8,710 participants with primary WGS data). For each participant, PTV and missense GLs were calculated separately as the sum of sHet values, as estimated by Sun et al.^13^, across all genes carrying at least one qualifying variant of that class, so that each affected gene contributed its selection coefficient once regardless of the number of qualifying variants it carried. The same sHet estimates were used in both cohorts. We estimated the correlation between PTV and missense GL in ABCD with Pearson’s *r* and a Fisher *z*-based 95% CI (Supplementary Figure S1).

### 4.6 Association models

Continuous outcomes were analyzed by ordinary least squares regression, fitted with statsmodels^66^ in Python. The quantitative phenotype and GL were each standardized within the phenotype-specific complete-case dataset, so coefficients represent standard deviations of outcome per standard deviation of GL. Incremental R^2^ was the difference between the full-model R^2^ and the R^2^ from the corresponding covariate-only model.

Binary outcomes were analyzed using a generalized linear model with binomial family and logit link. GL was standardized within the complete-case dataset, and exponentiated coefficients are odds ratios per standard-deviation increase in GL.

Both model classes used cluster-robust standard errors^67^ grouped by genetic family. In ABCD, families were defined by the ABCD genetic family identifiers, which group siblings and twins. In AoU, families were the connected components of the relatedness graph with pairwise kinship coefficient *≥* 0.1, which includes second-degree relatives. Participants without a family identifier were assigned participant-specific clusters. Base covariates were phenotype-appropriate age, sex, site and the first ten genetic ancestry principal components in both cohorts, together with Sentieon^68^ version in ABCD and sample source and sequencer in AoU. Sequencing covariates were log1p-transformed numbers of singletons, novel indels, filtered indels and total heterozygous depth, together with the proportion of GQ0 variants, heterozygous-to-homozygous-alternate ratio and SNP reference bias in ABCD, and log1p-transformed numbers of singletons and total indels, log1p-transformed mean coverage and the SNP heterozygous-to-homozygous-alternate ratio in AoU. Models in both cohorts were additionally adjusted for the number of rare synonymous variants carried. As a sensitivity analysis in ABCD, we refitted the models as linear (cognition) or generalized linear (binary outcomes) mixed models with a family random intercept, using lme4^69^ in R^70^, and retained the cluster-robust models because the linear mixed model gave a similar cognition estimate and the ASD model was degenerate.

Models were fitted only with at least 500 complete cases for continuous outcomes, at least 20 cases and 20 controls for binary outcomes, at least 20 participants with nonzero GL, and, for binary models, at least five exposed cases and five exposed controls.

#### Multiple testing

False discovery rate-adjusted *q*-values were computed with the Benjamini– Hochberg procedure^71^ separately for the 10 exploratory tests in ABCD and the 12 in AoU (depression, anxiety, bipolar disorder, ADHD and suicide attempt, plus height in AoU, each with PTV and missense GL). The primary outcomes (NIH Toolbox total cognition and ASD in ABCD, and ASD and schizophrenia in AoU), ID, joint models, meta-analyses and other additional analyses were not corrected for multiple testing and are reported with nominal *P* values.

#### Meta-analysis

ABCD and AoU estimates for ASD were combined by inverse-variance-weighted fixed-effect meta-analysis of the log odds ratios per typical impacted constrained gene, and heterogeneity was assessed with Cochran’s *Q*.

#### Marginal standardization

In ABCD, the adjusted ASD risk ratio and risk difference compared mean predicted probabilities from the logistic model with every participant’s GL set to the mean or to the mean plus one typical increment (below)^38^.

We translated per-standard-deviation GL effects to one typical constrained gene-level hit using observed GL increments associated with one damaging variant in a constrained gene in the ABCD cohort. The PTV increment was 0.221, defined as the mean sHet among observed PTVs in constrained genes (sHet >0.1). The missense increment, the mean sHet of damaging missense carrier calls in constrained genes, was 0.259. For continuous outcomes, the scaled coefficient was calculated as the per-standard-deviation coefficient multiplied by the raw constrained increment and divided by the raw GL standard deviation in the phenotype-specific analysis sample of each cohort, so that ABCD and AoU effects refer to the same raw GL increment. For binary outcomes, the same contrast was applied on the log-odds scale, and the scaled log odds ratio and its confidence limits were exponentiated. We used the raw increment rather than a common SD because random loss of rare PTV calls, which was more frequent in ABCD (Supplementary Note 3), reduces the GL SD but not the per-unit effect. AoU log ORs per raw sHet unit (from the known-gene split and the ancestry analyses) were multiplied by the raw increment.

### 4.7 Constraint groups and carrier analyses

GL was the exposure in all primary and exploratory association tests, and the carrier analyses below describe the risk associated with carrying a constrained PTV. We defined a constrained PTV as a qualifying PTV in a gene with sHet >0.1, and carriers as participants with at least one constrained PTV. In ABCD, the mean PTV GL of carriers exceeded that of non-carriers by 0.231 units, or 1.04 typical constrained genes. Participants were grouped by the sHet of the most constrained PTV they carried: no qualifying PTV, PTVs only in genes with sHet *≤* 0.1, and most constrained PTV with sHet 0.1–0.2, 0.2–0.4 or >0.4. In AoU, ORs for each constraint group, for any carrier, and for carrying one or two constrained PTVs were estimated relative to non-carriers (participants with no constrained PTV, including those with PTVs only in genes with sHet *≤* 0.1), using the GL models with carrier indicators in place of GL. In ABCD, mean NIH Toolbox total cognition was estimated in each constraint group, and ORs for low cognition were estimated for carriers relative to non-carriers. For each AoU outcome, we report the percentage of cases who carry a constrained PTV alongside the carrier percentage among all analyzed participants, and in ABCD the same quantity was computed for children with parent-reported ASD and for children in the bottom 10% of NIH Toolbox total cognition. Observed prevalences and percentages are given with 95% Wilson CIs. Attributable fractions were estimated by model-based standardization with all participants set to non-carriers, which for rare outcomes equals the percentage of cases who carry multiplied by (1 − 1/OR)^72–74^. Their 95% CIs came from 500 draws of the model coefficients in AoU and from simulating the case percentage and the carrier OR in ABCD, where sparse covariate cells made the joint covariance unstable. In AoU, attributable fractions were also split into the parts from carriers with a constrained PTV in an established gene (DDG2P, SFARI or the SCHEMA genes) and from carriers with constrained PTVs only in other genes.

We tested whether the linear sHet weighting holds across the constraint range by fitting per-unit GL effects jointly for the parts of GL contributed by genes with sHet *≤* 0.01, 0.01–0.1, 0.1–0.2, 0.2–0.4 and >0.4, and testing their equality with a Wald test.

### 4.8 Established disease genes

We split PTV GL into the part contributed by known disease genes and the part contributed by all other genes, fitted both parts jointly, and expressed each per typical impacted constrained gene. For ID, ASD and cognition, established genes were dominant developmental disorder genes in DDG2P^27^ with definitive, strong or moderate evidence, or genes with SFARI Gene^28^ score 1 or 2 or a syndromic annotation (1,593 genes, lists downloaded September 2026). Genes were matched across lists by HGNC symbol. For schizophrenia, established genes were the nine autosomal genes among the ten exome-wide significant genes of the SCHEMA consortium^7^ (the tenth, *GRIA3*, is X-linked). Equality of per-unit effects in established and other genes was tested with a Wald test, and GL excluding DDG2P and SFARI genes was additionally analyzed as a separate exposure.

### 4.9 Polygenic scores

PGS were computed as sums of effect-allele dosages weighted by per-variant weights: in ABCD from imputed genotypes, and in AoU from v9 short-read WGS genotypes at the ACAF-threshold sites, with missing calls set to twice the effect-allele frequency. The cognitive PGS was CogPGT^40^, constructed with SBayesRC^39^. The schizophrenia PGS was constructed with SBayesRC from a meta-analysis of the Psychiatric Genomics Consortium wave 3 GWAS^75^ and the FinnGen^76^ GWAS of schizoaffective disorder, and the bipolar disorder PGS from the public summary statistics of the Psychiatric Genomics Consortium GWAS^77^. The ASD PGS was constructed with SBayesR^78^ from the GWAS of Grove et al.^37^. None of the source GWAS included AoU or ABCD participants. PGS were standardized within each analysis sample. The matched PGS was the cognitive PGS for cognition in ABCD and for ID in AoU, and the disorder-specific PGS for all other outcomes (for ASD in ABCD, the ASD and cognitive PGS jointly).

We estimated the correlation between each GL and the matched PGS (in unrelated participants in AoU), and refitted each GL model with the matched PGS as a continuous covariate, reporting the change in the GL estimate. A GL × PGS product term tested for interaction on the log-odds scale (binary outcomes) or on the outcome scale (cognition). For carrier × PGS analyses, participants were cross-classified by carrier status and tertile of the matched PGS. ORs are relative to non-carriers in the lowest PGS tertile. In ABCD, unadjusted mean cognition was compared between carriers and non-carriers within each cognitive-PGS tertile.

### 4.10 Ancestry analyses

In AoU, GL was constructed with the same pipeline for participants of all genetic ancestry groups assigned by the AoU classifier^34^ (African, Admixed American, East Asian, European, Middle Eastern and South Asian). We used one reduced covariate set in every ancestry group: age, sex, 16 genetic ancestry principal components, log-transformed mean coverage, rare synonymous variant count, site and sample source, with standard errors clustered on kinship components. This set omitted the sequencer and the singleton, indel and heterozygosity covariates of the main models, the latter because they scale with genetic diversity and therefore differ systematically between ancestry groups. Fits were checked for convergence, degenerate standard errors and a ratio of robust to model-based standard errors outside 0.33–3, and failing fits were deemed non-estimable. GL effects were estimated per raw sHet unit, so that an increment has the same meaning in every ancestry, and expressed per typical impacted constrained gene. PGS effects were estimated per SD of the PGS within each ancestry group. Portability was summarized as the ratio of the African-ancestry to the European-ancestry log OR (or *β* for cognition), with a 95% CI from the delta method. For comparison with the PGS, GL portability is expressed per SD of GL within each ancestry group. Because GL varies more in African-ancestry participants, these ratios are slightly higher than the per-raw-unit ratios given in Supplementary Note 2. Heterogeneity across ancestry groups was assessed with Cochran’s *Q*. In ABCD, the corresponding comparison for NIH Toolbox total cognition used whole-blood participants assigned African or European ancestry with probability *≥* 0.90.

EHR-only schizophrenia was recorded in 4.1% of African-ancestry and 0.8% of European-ancestry participants with EHR data (Supplementary Note 2), consistent with the documented overdiagnosis of psychotic disorders in Black patients in the US^79^. Because EHR-based schizophrenia and bipolar diagnoses may thus be recorded with different accuracy across groups, we additionally used more specific definitions in all ancestry groups. For schizophrenia, cases required both an EHR diagnosis and self-report, and controls had answered the survey without reporting schizophrenia and had EHR data with no qualifying code. For bipolar disorder, cases were defined by self-report, and controls had answered the survey without reporting bipolar disorder. We also examined EHR-only definitions. PTV GL and the matched PGS were analyzed under the same definitions.

### 4.11 Within-family analyses

In AoU, full siblings were pairs of European-ancestry participants with a kinship coefficient of at least 0.177 and below 0.354 and an age difference of less than 12 years, excluding participants in any pair with kinship *≥* 0.354 (duplicates and monozygotic twins), and families were the connected components of the sibling-pair graph. We identified 4,229 full-sibling pairs in 3,883 families. For binary outcomes, within-sibship ORs were estimated by conditional logistic regression stratified by sibship among sibships discordant for the outcome, using the covariates of the population models that vary within sibships. Because individual psychiatric diagnoses had few discordant sibships, we also analyzed a composite of ASD, schizophrenia and bipolar disorder and a broader composite that also included depression, broad anxiety, ADHD and suicide attempt. Composite controls had EHR data and were not cases for any component, without a survey requirement. Height was analyzed with sibling fixed effects. Within-sibship analyses required at least 20 cases and at least 20 discordant sibships. Within-sibship estimates were compared with the population estimates with a Wald test of their difference.

In ABCD, within-family analyses of NIH Toolbox total cognition used 375 families of dizygotic twins and non-twin siblings, identified by ABCD genetic zygosity calls. Monozygotic twins were excluded from within-family estimation because they share their GL. We fitted sibling fixed-effects models and a Mundlak model that separates within-family and between-family associations by including the family-mean GL, and tested whether the two differed. We compared the within-family estimate with the full-cohort estimate using a family-cluster bootstrap. We estimated call sharing within dizygotic twin and sibling pairs and assessed call reproducibility in monozygotic twin pairs and technical replicates (Supplementary Note 3).

## Declarations

### Ethics

ABCD Study procedures were approved by the institutional review boards of the participating sites, with a central institutional review board at the University of California, San Diego. Parents provided written informed consent and children provided assent. ABCD data were used under an approved NBDC Data Use Certification. All of Us participants provided informed consent under protocols overseen by the All of Us Institutional Review Board. Analyses of de-identified All of Us data were conducted by registered researchers in the All of Us Researcher Workbench under the program’s data use agreement.

## Data availability

Data from the Adolescent Brain Cognitive Development™ (ABCD) Study were obtained from the NIH Brain Development Cohorts (NBDC) Data Hub under an approved Data Use Certification. Analyses used ABCD Data Release 7.0 (DOI: 10.82525/8f3w-5260). All of Us Research Program data were accessed by authorized users through the All of Us Researcher Workbench, and analyses used the Controlled Tier Dataset v9 (C2025Q4R6). Study-level summary statistics generated in this study are available from the authors upon reasonable request, subject to the applicable ABCD and All of Us data-use and dissemination requirements.

## Code availability

Analysis code is available from the corresponding author on request.

## Data Availability

Data from the Adolescent Brain Cognitive Development (ABCD) Study were obtained from the NIH Brain Development Cohorts (NBDC) Data Hub under an approved Data Use Certification. Analyses used ABCD Data Release 7.0 (DOI: 10.82525/8f3w-5260). All of Us Research Program data were accessed by authorized users through the All of Us Researcher Workbench, and analyses used the Controlled Tier Dataset v9 (C2025Q4R6). Study-level summary statistics generated in this study are available from the authors upon reasonable request, subject to the applicable ABCD and All of Us data-use and dissemination requirements.

## Acknowledgements.

Data used in the preparation of this article were obtained from the Adolescent Brain Cognitive Development™ (ABCD) Study, held in the NIH Brain Development Cohorts Data Sharing Platform. This is a multisite, longitudinal study designed to recruit more than 10,000 children aged 9–10 and follow them over 10 years into early adulthood. The ABCD Study® is supported by the National Institutes of Health and additional federal partners under award numbers U01DA041048, U01DA050989, U01DA051016, U01DA041022, U01DA051018, U01DA051037, U01DA050987, U01DA041174, U01DA041106, U01DA041117, U01DA041028, U01DA041134, U01DA050988, U01DA051039, U01DA041156, U01DA041025, U01DA041120, U01DA051038, U01DA041148, U01DA041093, U01DA041089, U24DA041123 and U24DA041147.

A full list of supporters is available at https://abcdstudy.org/federal-partners/. ABCD Consortium investigators designed and implemented the study and/or provided data but did not necessarily participate in the analysis or writing of this report. This manuscript reflects the views of the authors and may not reflect the opinions or views of the NIH or ABCD Consortium investigators. The ABCD data repository grows and changes over time. The ABCD data used in this report came from NBDC Digital Object Identifier (DOI) 10.82525/8f3w-5260.

We gratefully acknowledge All of Us participants for their contributions, without whom this research would not have been possible. We also thank the National Institutes of Health’s All of Us Research Program for making available the participant data examined in this study.

## Funding

This work was funded by Herasight, Inc.

## Competing interests

M.A., S.M., J.S., J.A. and T.W. are employees of Herasight, Inc. and hold equity in the company. A.S.Y. is an advisor to Herasight, Inc.

## Author contributions

Mohammad Ahangari: Conceptualization, Formal analysis, Writing – review & editing

Spencer Moore: Formal analysis, Writing – review & editing

Justin Schleede: Conceptualization

Jonathan Anomaly: Conceptualization, Writing – review & editing

Alexander Strudwick Young: Conceptualization, Writing – review & editing

Tobias Wolfram: Conceptualization, Formal analysis, Investigation, Methodology, Project administration, Supervision, Visualization, Writing – original draft, Writing – review & editing

## Supplementary material

**Figure S1:**
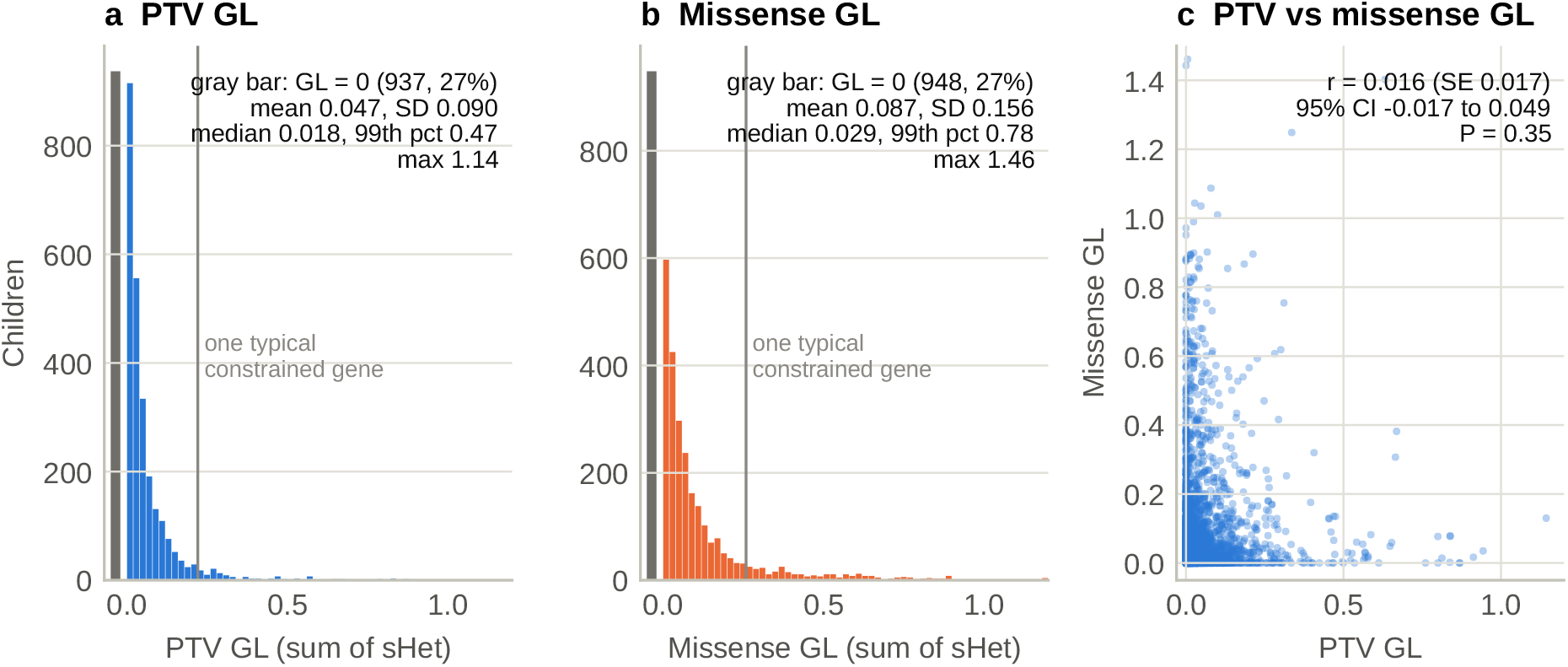
Distributions of PTV and missense GL in ABCD. (a) PTV GL and (b) damaging missense GL (sum of sHet) in 3,523 European-ancestry children. PTV GL had a mean of 0.047 and an SD of 0.090, with 26.6% of children at zero and a 99th percentile of 0.47. Missense GL had a mean of 0.087 and an SD of 0.156, with 26.9% at zero and a 99th percentile of 0.78. Gray bars show a GL of zero, and values above 1.2 are pooled into the last bin. Vertical lines mark one typical impacted constrained gene (0.221 for PTV and 0.259 for missense GL). Many of the 8.9% of children who carry a constrained PTV lie to the left of the PTV line, because 62% of carriers have their most constrained PTV in a gene with sHet of 0.1–0.2, below the typical increment. (c) PTV GL against missense GL (r = 0.016). In AoU (302,421 participants), PTV GL had a mean of 0.062 and an SD of 0.103, with 21.0% at zero and a 99th percentile of 0.54, and missense GL had a mean of 0.081 and an SD of 0.151, with 32.5% at zero and a 99th percentile of 0.77.

**Figure S2:**
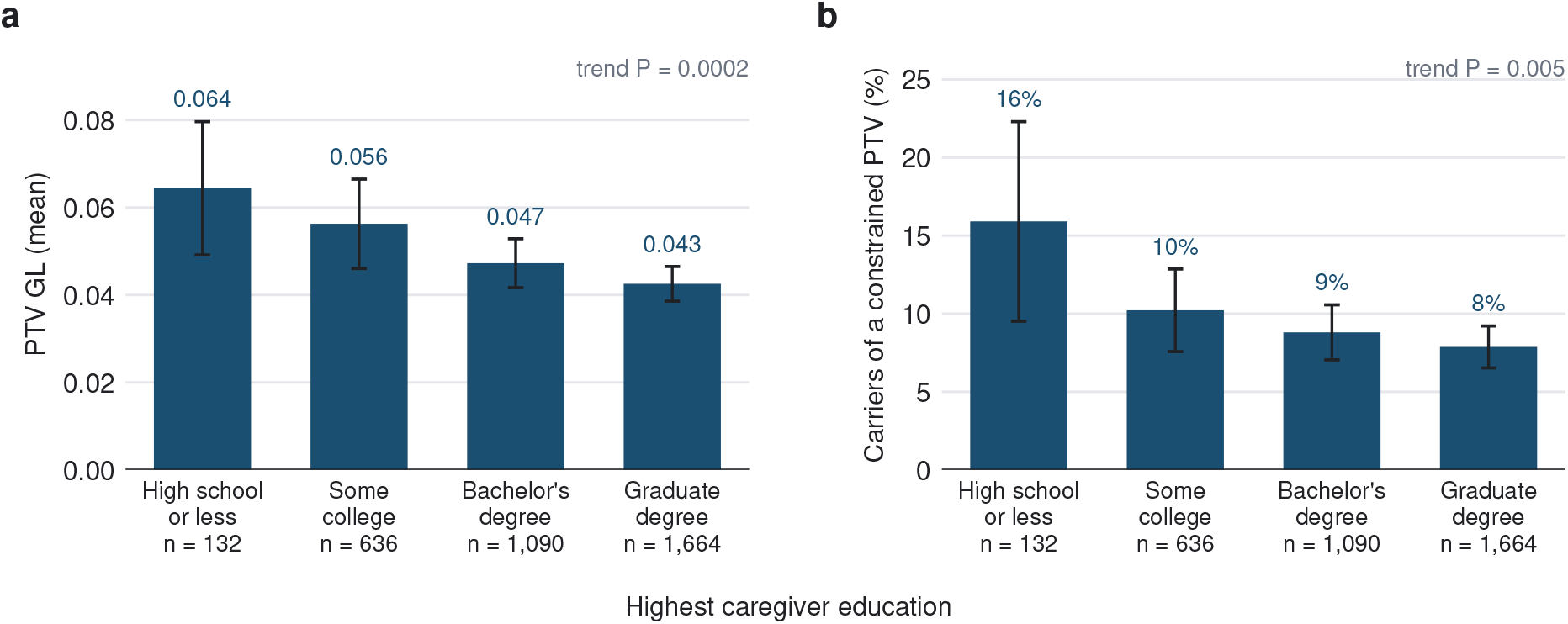
PTV GL by caregiver education in ABCD. (a) Mean PTV GL and (b) percentage of children carrying a constrained PTV, by the highest education of either caregiver (3,522 children, with the 22 children of caregivers without a high-school diploma pooled into the high-school group). Bars show means with 95% CIs clustered by family. Trend *P* values are for a linear term in education level, adjusted for the covariates of the primary models. Household income showed a weaker gradient (trend *P* = 0.03 for mean PTV GL and 0.6 for carrier status).

### Supplementary Notes

**Supplementary Note 1: Calibration of the sHet weighting.** We split PTV GL in AoU into five sHet bins and estimated their per-unit effects in a joint model (Methods). Equal per-unit effects were rejected for schizophrenia (*P* = 2×10^−4^), bipolar disorder (*P* = 1×10^−5^), depression (*P* = 9×10^−6^) and anxiety (*P* = 8×10^−6^), nominally for ASD (*P* = 0.012) and ADHD (*P* = 0.013), and not for intellectual disability (*P* = 0.14) or suicide attempt (*P* = 0.07). For the psychiatric outcomes other than suicide attempt, per-unit effects were largest at sHet 0.1–0.2 and, for ASD and schizophrenia, about half as large above 0.4 on the log-odds scale. Intellectual disability showed the same pattern without significant heterogeneity (2.92 per typical gene at sHet 0.1–0.2 and 1.91 above 0.4). Carrier ORs nevertheless increased with constraint for every outcome (Figure 2c), because carriers in the most constrained genes carry several times more sHet units than carriers in moderately constrained genes, which outweighs the smaller effect per unit. Besides the fact that sHet indexes total fitness cost rather than the effect on a specific outcome (Discussion), possible explanations include under-representation of carriers of the most damaging PTVs among adult volunteers and more false-positive or nonsense-mediated decay-escaping PTV calls in the most constrained genes.

**Supplementary Note 2: Phenotype definitions and the ancestry comparison.** Under the main AoU definitions, participants without survey data could be cases but not controls. Definitions based on EHR diagnoses alone, for which control status does not depend on survey completion, gave European-ancestry associations of 90% (schizophrenia), 84% (bipolar disorder) and 101% (ASD) of those under the main definitions on the log-odds scale. The ASD association was also consistent across ancestries when ASD was defined from EHR diagnoses alone, a definition that does not require survey completion (heterogeneity *P* = 0.73). Under the main definitions, the African-ancestry schizophrenia and bipolar disorder associations were about half the European-ancestry associations on the log-odds scale (heterogeneity *P* < 10^−5^ and *P* = 3×10^−4^). These definitions draw cases from any qualifying EHR code but require controls to have completed the relevant survey, which 29% of African-ancestry and 63% of European-ancestry participants had done. EHR-only definitions remained attenuated (schizophrenia *P* = 7×10^−5^, bipolar disorder *P* = 8×10^−6^), and EHR-only schizophrenia prevalence was 4.1% in African-ancestry and 0.8% in European-ancestry participants with EHR data, consistent with diagnostic misclassification. Under more specific definitions (schizophrenia requiring both an EHR diagnosis and self-report, and self-reported bipolar disorder), the PTV GL associations were consistent across ancestries. Per raw sHet unit, the African-ancestry PTV GL effect was 78% of the European-ancestry effect for intellectual disability (*P* = 0.02 for the difference), 116% for ASD, 89% for schizophrenia (heterogeneity *P* = 0.30), 66% for bipolar disorder (heterogeneity *P* = 0.40) and 90% for cognition in ABCD (Figure 4b shows the corresponding per-SD values).

**Supplementary Note 3: Call reproducibility and GL sharing within families.** In ABCD, siblings shared 43% of PTV calls and 48% of missense calls, consistent with the Mendelian expectation of 50% once call reproducibility is taken into account (50% × 85% *≈* 43% for PTVs). Genetically identical co-twins shared 99.8% of missense calls but only 85% of PTV calls (95% of SNV and 76% of frameshift-indel PTVs), most likely reflecting false-negative indel calls under the stricter indel genotype filters. Duplicate sequencing of 130 children implied a GL reliability of 0.96 in the full cohort and 0.94 within families, so calling noise adds only about 2% to the attenuation of within-family estimates.

In AoU, 4,229 full-sibling pairs shared 46.7% of each other’s qualifying PTV calls and 46.5% of missense calls, and 1,793 monozygotic-twin and duplicate pairs shared 96.7% and 97.8%, indicating a call reproducibility of approximately 97%. Constrained PTVs (sHet > 0.1) were shared less often (44.4% of 1,089 calls). Relative to qualifying PTVs in the least constrained genes (sHet *≤* 0.01, 47.7%), this implies that about 93% (95% CI 84–103%) were inherited and that the remaining 7% arose *de novo* or were false-positive calls, with a larger but imprecise share for sHet > 0.4 (17%, 95% CI −9 to 45%). When sibling GLs correlate at 0.5, the variance of the within-pair GL difference is expected to equal the population GL variance. The ratio of the two was 1.06 for PTV GL. PTV GL therefore varies substantially within families, which allows associations to be estimated within sibships.

---

**Table S1:** Association results for exome-wide rare variant genetic load. Each row reports one regression of an outcome on a protein-truncating (PTV) or damaging missense genetic load (GL) in ABCD, All of Us, or their fixed-effect meta-analysis. Marginal models include one GL plus covariates, whereas joint models include the PTV and missense GLs simultaneously. Effects per SD of GL are standardized regression coefficients (*β*, SD of outcome per SD of GL) for continuous outcomes and odds ratios (OR) for binary outcomes, with 95% confidence intervals from cluster-robust standard errors. Effects per typical gene rescale the per-SD effect to one damaging variant in a typical impacted constrained gene (heterozygous selection coefficient, sHet, > 0.1), multiplying by 0.221 (PTV) or 0.259 (missense) and dividing by the GL SD of the analysis sample (Methods). For the meta-analysis rows, *P* refers to the meta-analysis per typical gene (Methods). *q* values are Benjamini–Hochberg false discovery rate-adjusted *P* values for each cohort’s exploratory marginal models (Methods) and were not computed for the primary outcomes, intellectual disability, joint models or meta-analyses (–). ASD, autism spectrum disorder.

| Cohort | Outcome | Model | GL | n | Cases/controls | Scale | Effect per SD of GL (95% CI) |  | P | q | Effect per typical gene (95% CI) |  |
| --- | --- | --- | --- | --- | --- | --- | --- | --- | --- | --- | --- | --- |
| ABCD | NIH total cognition | Marginal | PTV | 3,498 | – | $\beta$ | –0.088 | (–0.118, –0.059) | $4.5 \times 10^{-9}$ | – | –0.218 | (–0.291, –0.145) |
| ABCD | NIH total cognition | Joint | PTV | 3,498 | – | $\beta$ | –0.088 | (–0.117, –0.058) | $7.3 \times 10^{-9}$ | – | –0.216 | (–0.290, –0.143) |
| ABCD | NIH total cognition | Marginal | Missense | 3,498 | – | $\beta$ | –0.044 | (–0.079, –0.010) | 0.011 | – | –0.074 | (–0.130, –0.017) |
| ABCD | NIH total cognition | Joint | Missense | 3,498 | – | $\beta$ | –0.043 | (–0.078, –0.009) | 0.015 | – | –0.072 | (–0.129, –0.014) |
| ABCD | ASD | Marginal | PTV | 3,506 | 46/3,460 | OR | 1.27 | (1.04, 1.55) | 0.017 | – | 1.80 | (1.11, 2.92) |
| All of Us | ASD | Marginal | PTV | 188,189 | 2,797/185,392 | OR | 1.14 | (1.11, 1.17) | $7.8 \times 10^{-22}$ | – | 1.34 | (1.26, 1.42) |
| Meta-analysis | ASD | FE meta | PTV | 191,695 | 2,843/188,852 | OR | 1.14 | (1.11, 1.17) | $9.0 \times 10^{-23}$ | – | 1.35 | (1.27, 1.43) |
| ABCD | ASD | Marginal | Missense | 3,506 | 46/3,460 | OR | 1.24 | (0.99, 1.56) | 0.061 | – | 1.44 | (0.98, 2.11) |
| All of Us | ASD | Marginal | Missense | 188,189 | 2,797/185,392 | OR | 1.06 | (1.02, 1.10) | 0.0011 | – | 1.10 | (1.04, 1.17) |
| Meta-analysis | ASD | FE meta | Missense | 191,695 | 2,843/188,852 | OR | 1.06 | (1.03, 1.10) | $4.3 \times 10^{-4}$ | – | 1.11 | (1.05, 1.18) |
| All of Us | Schizophrenia | Marginal | PTV | 189,009 | 2,306/186,703 | OR | 1.20 | (1.17, 1.23) | $3.5 \times 10^{-45}$ | – | 1.51 | (1.42, 1.59) |
| All of Us | Schizophrenia | Marginal | Missense | 189,009 | 2,306/186,703 | OR | 1.08 | (1.04, 1.12) | $2.2 \times 10^{-5}$ | – | 1.15 | (1.08, 1.22) |
| All of Us | Intellectual disability | Marginal | PTV | 225,299 | 396/224,903 | OR | 1.39 | (1.33, 1.44) | $4.0 \times 10^{-53}$ | – | 2.02 | (1.85, 2.21) |
| All of Us | Intellectual disability | Marginal | Missense | 225,299 | 396/224,903 | OR | 1.15 | (1.07, 1.23) | $1.7 \times 10^{-4}$ | – | 1.26 | (1.12, 1.42) |
| ABCD | Depression | Marginal | PTV | 3,523 | 722/2,801 | OR | 1.11 | (1.02, 1.20) | 0.012 | 0.12 | 1.28 | (1.05, 1.56) |
| All of Us | Depression | Marginal | PTV | 215,863 | 108,279/107,584 | OR | 1.07 | (1.06, 1.08) | $6.0 \times 10^{-50}$ | $2.4 \times 10^{-49}$ | 1.16 | (1.14, 1.18) |
| ABCD | Depression | Marginal | Missense | 3,523 | 722/2,801 | OR | 1.04 | (0.95, 1.13) | 0.44 | 0.55 | 1.06 | (0.91, 1.23) |
| All of Us | Depression | Marginal | Missense | 215,863 | 108,279/107,584 | OR | 1.02 | (1.01, 1.03) | $1.3 \times 10^{-6}$ | $1.6 \times 10^{-6}$ | 1.04 | (1.02, 1.05) |
| ABCD | Anxiety disorder | Marginal | PTV | 3,523 | 684/2,839 | OR | 1.06 | (0.98, 1.14) | 0.17 | 0.38 | 1.14 | (0.94, 1.38) |
| All of Us | Anxiety disorder | Marginal | PTV | 217,566 | 102,607/114,959 | OR | 1.07 | (1.06, 1.08) | $7.8 \times 10^{-45}$ | $2.3 \times 10^{-44}$ | 1.15 | (1.13, 1.17) |
| ABCD | Anxiety disorder | Marginal | Missense | 3,523 | 684/2,839 | OR | 1.04 | (0.95, 1.13) | 0.41 | 0.55 | 1.06 | (0.92, 1.22) |
| All of Us | Anxiety disorder | Marginal | Missense | 217,566 | 102,607/114,959 | OR | 1.03 | (1.02, 1.04) | $3.1 \times 10^{-10}$ | $5.3 \times 10^{-10}$ | 1.05 | (1.03, 1.07) |
| ABCD | Bipolar disorder | Marginal | PTV | 3,523 | 264/3,259 | OR | 0.95 | (0.84, 1.08) | 0.42 | 0.55 | 0.88 | (0.65, 1.20) |
| All of Us | Bipolar disorder | Marginal | PTV | 193,529 | 15,990/177,539 | OR | 1.13 | (1.12, 1.15) | $2.9 \times 10^{-80}$ | $3.5 \times 10^{-79}$ | 1.32 | (1.28, 1.36) |
| ABCD | Bipolar disorder | Marginal | Missense | 3,523 | 264/3,259 | OR | 1.03 | (0.91, 1.15) | 0.68 | 0.75 | 1.04 | (0.86, 1.27) |
| All of Us | Bipolar disorder | Marginal | Missense | 193,529 | 15,990/177,539 | OR | 1.04 | (1.02, 1.06) | $5.6 \times 10^{-7}$ | $7.5 \times 10^{-7}$ | 1.07 | (1.04, 1.10) |

| Cohort | Outcome | Model | GL | n | Cases/controls | Scale | Effect per SD of GL (95% CI) | P | q | Effect per typical gene (95% CI) |
| --- | --- | --- | --- | --- | --- | --- | --- | --- | --- | --- |
| ABCD | ADHD | Marginal | PTV | 3,523 | 1,037/2,486 | OR | 1.07 (0.99, 1.15) | 0.083 | 0.28 | 1.18 (0.98, 1.42) |
| All of Us | ADHD | Marginal | PTV | 192,412 | 24,944/167,468 | OR | 1.04 (1.03, 1.05) | $8.9 \times 10^{-9}$ | $1.3 \times 10^{-8}$ | 1.09 (1.06, 1.12) |
| ABCD | ADHD | Marginal | Missense | 3,523 | 1,037/2,486 | OR | 1.07 (0.99, 1.15) | 0.082 | 0.28 | 1.12 (0.99, 1.26) |
| All of Us | ADHD | Marginal | Missense | 192,412 | 24,944/167,468 | OR | 1.01 (0.99, 1.02) | 0.25 | 0.25 | 1.01 (0.99, 1.04) |
| ABCD | Suicide attempt | Marginal | PTV | 3,523 | 151/3,372 | OR | 1.09 (0.96, 1.24) | 0.19 | 0.38 | 1.24 (0.90, 1.71) |
| All of Us | Suicide attempt | Marginal | PTV | 91,497 | 9,248/82,249 | OR | 1.08 (1.06, 1.10) | $7.6 \times 10^{-15}$ | $1.5 \times 10^{-14}$ | 1.19 (1.14, 1.25) |
| ABCD | Suicide attempt | Marginal | Missense | 3,523 | 151/3,372 | OR | 0.97 (0.78, 1.20) | 0.78 | 0.78 | 0.95 (0.67, 1.36) |
| All of Us | Suicide attempt | Marginal | Missense | 91,497 | 9,248/82,249 | OR | 1.03 (1.01, 1.05) | 0.010 | 0.011 | 1.05 (1.01, 1.09) |
| All of Us | Height | Marginal | PTV | 292,676 | - | $\beta$ | -0.026 (-0.029, -0.023) | $1.0 \times 10^{-74}$ | $6.0 \times 10^{-74}$ | -0.056 (-0.062, -0.050) |
| All of Us | Height | Marginal | Missense | 292,676 | - | $\beta$ | -0.012 (-0.014, -0.009) | $7.9 \times 10^{-17}$ | $1.9 \times 10^{-16}$ | -0.020 (-0.025, -0.015) |

**Table S2:** Population and within-sibship associations. Values are per typical impacted constrained gene (ORs for binary outcomes, SD for height and cognition), with scaling and population estimates as in Table 1. For binary outcomes, within-sibship ORs come from conditional logistic regression on AoU sibships discordant for the outcome, whereas height (AoU) and NIH total cognition (ABCD) use sibling fixed-effects models. Models for ASD and schizophrenia did not converge, and intellectual disability had fewer than 20 cases among siblings. *P* (PTV) tests within-sibship against population estimates, for ABCD cognition with a family-cluster bootstrap. The ABCD within-family estimate differed nominally from the between-family estimate in the same 375 families (*P* = 0.04), which was unusually steep (−0.35 versus −0.22 SD in the full cohort).

| Outcome | Cohort | Sibships | PTV: population | PTV: within sibships | P (within vs population, PTV) | Missense: population | Missense: within sibships |
| --- | --- | --- | --- | --- | --- | --- | --- |
| ASD, schizophrenia or bipolar disorder | AoU | 255 discordant | 1.27 (1.24–1.31) | 1.83 (1.11–3.01) | 0.16 | 1.06 (1.03–1.09) | 0.79 (0.53–1.18) |
| Bipolar disorder | AoU | 168 discordant | 1.32 (1.28–1.36) | 1.32 (0.70–2.48) | 0.99 | 1.07 (1.04–1.10) | 1.04 (0.55–1.97) |
| Depression | AoU | 915 discordant | 1.16 (1.14–1.18) | 1.12 (0.82–1.53) | 0.85 | 1.04 (1.02–1.05) | 1.13 (0.91–1.42) |
| Anxiety | AoU | 879 discordant | 1.15 (1.13–1.17) | 0.93 (0.66–1.33) | 0.24 | 1.05 (1.03–1.07) | 1.05 (0.84–1.31) |
| ADHD | AoU | 287 discordant | 1.09 (1.06–1.12) | 1.11 (0.55–2.26) | 0.94 | 1.01 (0.99–1.04) | 1.25 (0.84–1.87) |
| Suicide attempt | AoU | 72 discordant | 1.19 (1.14–1.25) | 1.27 (0.35–4.56) | 0.93 | 1.05 (1.01–1.09) | 0.55 (0.08–3.99) |
| Any of seven conditions | AoU | 1,123 discordant | 1.09 (1.07–1.11) | 1.20 (0.92–1.55) | 0.50 | 1.02 (1.01–1.04) | 1.07 (0.89–1.30) |
| Height (SD) | AoU | 3,711 | −0.056 (−0.062 to −0.050) | −0.076 (−0.139 to −0.014) | 0.52 | −0.020 (−0.025 to −0.015) | 0.002 (−0.034 to 0.038) |
| NIH total cognition (SD) | ABCD | 375 | −0.218 (−0.291 to −0.145) | −0.085 (−0.255 to 0.085) | 0.16 | −0.074 (−0.130 to −0.017) | 0.005 (−0.234 to 0.245) |

## Notes

### Author Declarations

The Institutional Review Board of the University of California, San Diego (central IRB of the Adolescent Brain Cognitive Development Study) gave ethical approval for the ABCD Study, and the All of Us Institutional Review Board gave ethical approval for the All of Us Research Program. This work analyzed de-identified ABCD data under an approved NIH Brain Development Cohorts Data Use Certification and de-identified All of Us Controlled Tier data in the All of Us Researcher Workbench under the program's data use agreement.

